# Integrative molecular profiling identifies two molecularly and clinically distinct subtypes of blastic plasmacytoid dendritic cell neoplasm

**DOI:** 10.1101/2022.05.26.22275640

**Authors:** Axel Künstner, Julian Schwarting, Hanno M. Witte, Veronica Bernard, Stephanie Stölting, Kathrin Kusch, Kumar Nagarathinam, Nikolas von Bubnoff, Eva M. Murga Penas, Hartmut Merz, Hauke Busch, Alfred C. Feller, Niklas Gebauer

**Affiliations:** University Cancer Center Schleswig-Holstein, University Hospital of Schleswig-Holstein, Campus Lübeck, 23538 Lübeck, Germany; Medical Systems Biology Group, University of Lübeck, Ratzeburger Allee 160, 23538 Lübeck, Germany; Institute for Cardiogenetics, University of Lübeck, Ratzeburger Allee 160, 23538 Lübeck, Germany; Department of Hematology and Oncology, University Hospital of Schleswig-Holstein, Campus Lübeck, Ratzeburger Allee 160, 23538 Lübeck, Germany; Department of Hematology and Oncology, Federal Armed Forces Hospital Ulm, Oberer Eselsberg 40, 89081 Ulm; Hämatopathologie Lübeck, Reference Centre for Lymph Node Pathology and Hematopathology, Maria-Goeppert-Straße 9a, 23562 Lübeck, Germany; Institute of Biochemistry, University of Lübeck, Ratzeburger Allee 160, 23538 Lübeck, Germany; Institute of Human Genetics, University Hospital of Schleswig-Holstein, Campus Kiel, Schwanenweg 24, 24105 Kiel, Germany

**Keywords:** blastic plasmacytoid dendritic cell neoplasm, whole-exome sequencing, RNA-sequencing, RRBS

## Abstract

Blastic plasmacytoid dendritic cell neoplasm (BPDCN) is an aggressive malignancy assumed to originate from plasmacytoid dendritic cells (pDCs), which mostly affects the skin, bone marrow, lymph nodes and sequentially other organ systems. RNA-, targeted- and exome sequencing studies have identified molecular characteristics, associated with BPDCN-pathogenesis, yet an integrative molecular assessment of BPDCN remains pending. Here, we combined paired WES/RNA-Seq with genome-wide copy-number analysis to characterize 47 BPDCN patients for mutational drivers, cytogenetic aberrations and gene-expression profiles. We identified alterations in epigenetic regulators (*TET2, EP300, DNMT3A, SF3B1, EZH2*) and a mutational disruption of RTK-RAS signaling (*NF1, NRAS, EGFR*) as drivers of BPDCN alongside deletions of tumor suppressors (*CDKN2A, RB1, TP53*), amplifications of oncogenes (*IDH2, MET, EZH2*) and recurrent fusions (*MYB, ALK*). The mutational landscape further provides evidence for frequent induction of PDGF signaling and extracellular matrix interactions as well as a gender specificity and a subset of MSI^high^ patients. Many genes affected in BPDCN are shared with chronic myelomonocytic leukemia (CMML), emphasizing a close relationship between these entities and to a lesser extent with acute myeloid leukemia (AML). Ontological assessment of RNA-Seq data revealed two BPDCN subtypes, a typical pDC-derived subtype (C1) and a (common) cDC-enriched subtype (C2), which were then shown to exhibit distinct mutational (*EP300, ARID2, NF1* mutations in typical pDC vs. *DNMT3A, SRSF2* mutations in the cDC-enriched subtype) and clinical features.

In summary, our hitherto most comprehensive characterization of BPDCN reveals molecular hallmarks alongside actionable vulnerabilities and highlights two novel subtypes that are molecularly and clinically distinct.

**Key Points:**

- Paired WES/RNA-Seq and copy number analysis of a large BPDCN cohort reveals two molecularly and clinically distinct subtypes.
- Multi-omics identify recurrent therapeutic targets and vulnerabilities including MSI^high^ and mutations within epigenetic regulation of gene expression and RTK-RAS signaling.

## Introduction

Blastic plasmacytoid dendritic cell neoplasm (BPDCN) is a rare (approx. 0.5% of hematological malignancies) and aggressive blood cancer. Initially considered to be a blastic NK-cell lymphoma, the discovery of its predominant cellular descent from non-activated, CD56^+^ plasmacytoid dendritic cells (pDC) has led to the recognition of BPDCN as an independent entity within the WHO classification of myeloid neoplasms ^1,2^. Cutaneous involvement often precedes bone marrow infiltration and dissemination into lymph nodes and extranodal organs. Some patients have primarily leukemic presentation with bone marrow involvement. Median age at diagnosis lies within the seventh decennium and a male predominance (4:1) is observed ^3^. The specific immunophenotype of CD4^+^, CD56^+^, and CD123^+^ reliably enables specific diagnosis although, differential diagnosis between BPDCN and acute myeloid leukemia (AML) can be challenging. Co-expression of CD303 and further pDC–associated antigens (e.g. TCL1) may aid in the delineation from AML, while expression of B-(CD79b), T-(CD2, CD7) and precursor antigens (TdT) occur ^4^. Recent observations proposed a subset of BPDCN, including the CAL-1 cell line, to originate from AXL1^+^ SIGLEC6^+^ DCs (AS-DCs), which suggests a diverse cellular ontogeny ^5,6^. Prognosis was poor in the era of conventional cytotoxic chemotherapy with high rates of relapse and primary refractory disease. Acute leukemia-derived protocols appear to be superior to non-Hodgkin lymphoma (NHL)-like approaches resulting in a median overall survival (OS) of 12 to 14 months. Usually, intensive consolidation as either allogenic or autologous stem cell transplantation is recommended ^3,7,8^. Correspondingly, therapy with curative intent was reserved for young and otherwise healthy patients, up until the introduction of tagraxofusp, a CD123-directed cytotoxin, which recently demonstrated high clinical efficacy in a phase 1/2 trial, including elderly patients ^9^. Whether these initial results translate into long-turn survival benefits, especially in patients unfit for intensive consolidation, remains questionable.

A genetically defined taxonomy is already embedded in the WHO classification ^1^. Given molecularly tailored treatments increasingly outperforming non-biomarker stratified therapies in patients with advanced cancers, an entity-specific in-depth molecular definition has become a vital prerequisite in precision oncology ^10^. Panel or whole-exome sequencing (WES) studies on small cohorts of patients and RNA-sequencing (RNA-Seq) of selected cases have reported a limited number of potential genetic drivers and transcriptional mechanisms in BPDCN ^5,11-18^. Molecular and pathogenetic similarities between BPDCN and several other blood cancers have been proposed, as both syn- and metachronous myeloid neoplasms such as chronic myelomonocytic leukemia (CMML), AML and even myelodysplastic syndromes (MDS) have been reported in up to 20% of cases ^19^. This is further reflected in the mutational features of BPDCN, comprising mutations in epigenetic regulation (*TET2, ASXL1, EZH2*), RAS signaling (*NRAS, KRAS*), splicing (*ZRSR2, SF3B1*) and tumor suppressors (TSGs; *TP53, ATM*). Recently, mutational disruption of epigenetic regulators except RTK-RAS and TSGs was shown to be a recurrent feature of clonal hematopoiesis, frequently underlying BPDCN ^20^. A comprehensive and integrative molecular characterization of a representative and clinically annotated cohort of BPDCN patients has not been performed so far. In this study, we combined paired WES and RNA-Seq with genome-wide copy number analysis of 47 cases. Through this integrated analysis, we refine the position of BPDCN within the molecular spectrum of hematological malignancies and describe two novel subtypes that are transcriptionally, genomically and clinically distinct offering the possibility for novel, personalized therapy approaches.

## Materials and methods

### Case selection, extraction of nucleic acids, WES and RNA-Seq

For this retrospective analysis, we reviewed our institutional archive for cases of histologically confirmed BPDCN between January 2001 and December 2020. The study was approved by the ethics committee of the University of Lübeck (reference-no 18-311) and conducted in accordance with the declaration of Helsinki. Genomic DNA and RNA were extracted from three 5µm FFPE tissue sections of either tumor or normal tissue (where available; n = 3) employing Maxwell® RSC DNA FFPE kit and Maxwell® RSC RNA FFPE kit (both Promega). WES and RNA-Seq following library preparation using Agilent SureSelect Human All Exon V6 library preparation kit (Agilent Technologies) and NEBNext® UltraT Directional RNA Library Prep Kit (New England BioLabs), respectively were performed on a NovaSeq platform (Illumina) at Novogene (UK) Co. as described ^21^. Tumor whole exome libraries were sequenced to a median depth of 119x (mean 129 ± 54 s.d.) and normal libraries reached a median depth of 67x (mean 83 ± 36 s.d.).

### Exome Data Processing and Variant Calling

Raw sequencing data (fastq format) was trimmed (adapter and quality values) applying FASTP_22_ (v0.23.0; minimum length, 50 bp; maximum unqualified bases, 30%; trim tail set to 1), trimmed reads were mapped to GRCh38 using BWA MEM (v0.7.15)^23^ and mappings were converted into *BAM* format using Picard Tools (v2.18.4)^24^. Afterwards, mate-pair information was fixed, PCR duplicates were removed, and base quality recalibration was perform using PICARD TOOLS, GATK (v4.2.3.0)^25^ and dbSNP v138^26^. Single nucleotide variants (SNVs) and short insertions and deletions (indels) were identified following GATKs best practices for somatic mutation calling. A more detailed description of the variant calling is given in the Supplementary Material and Methods.

Selected mutations in *TET2, IDH1, RUNX1, ASXL1* and *TP53* in samples from which sufficient DNA was available for confirmatory investigations were subjected to either Sanger sequencing or amplicon based NGS which allowed confirmation of all investigated variants (**Supplementary Table 2**).

### RNA-Seq Data Processing, Quantification, Fusion Detection, and Deconvolution

Gene expression profiles were retrieved from adapter trimmed reads (FASTP v0.23.0) using STAR aligner (v2.7.4b) against GRCh38 (GENCODE v37) as reference ^27^. Differentially expressed genes on normalized expression values between two conditions were identified using a linear modelling approach (LIMMA package, v3.50.1)^28^ and pathway enrichment was performed using a rank-MANOVA based approach. Fusions were detected using FUSIONCATCHER (v1.33)^29^. Further details are given in the Supplementary Material and Methods.

Previously published scRNA-data from Villani *et al*. was used to infer the cell-type composition of the bulk RNA-Seq data with respect to dendritic cells and monocytes applying a deconvolution of damped weighted least squares (DWLS) method. Briefly, a signature matrix was built from the single cell data using default parameters (diff.cutoff = 0.5, pval.cutoff=0.01) and deconvolution was performed applying the *solveDampenedWLS* method^30,31^. The optimal number of clusters was inferred applying the average silhouette method (FACTOEXTRA package v1.0.7) on deconvolution values.

### Detection of Somatic Copy Number Alterations

Somatic copy number alterations were detected using OncoScan CNV assays (ThermoFisher). Raw data (*CEL* files) were processed using the EACON package (v0.3.6-2) with SEQUENZA^32^ as segmentation algorithm. *L2R* files in CBS format were used as input for GISTIC (v2.0.22)^33^ to identify regions that are significantly amplified or deleted across all samples (confidence level 0.95, focal length cutoff 0.5, q-value threshold 0.1). Impact of identified regions on pathogenicity was inferred using X-CNV^34^ with standard parameters.

## Statistical analysis

If not stated differently, statistical analysis was performed using R (v4.1.2) and p-values were corrected using Benjamini-Hochberg correction. For further details see Supplementary Material and Methods.

## Results

### Clinical characteristics of the study group

From initially 74 patients we collected 47 cases of BPDCN at diagnosis with sufficient FFPE tissue samples for molecular studies (mean/median age 69.0/74.0 years; range 15 – 91 years) all of which were included in the final analysis. The majority of patients in our study were male (34/47; 72%) and presented with cutaneous and other extranodal involvement. Of primary responders, 22% went on to autologous/allogeneic stem cell transplantation. Median progression-free and overall survival was 8 and 12 months, respectively. Best supportive care was provided for four (15%) elderly/frail patients who refused any type of anticancer treatment and rapidly succumbed to BPDCN progression. Baseline clinicopathological characteristics of BPDCN cases included in the current study are briefly summarized in **Supplementary Table 3** and **Table 1**.

**Table 1.** Baseline clinicopathological characteristics of the study group

| Characteristics | BPDCN<br>(n = 47) |
| --- | --- |
| Age (yrs.; median (range)) | 74 (15 - 91) |
| **Sex** |  |
| Female | 13/47 (28%) |
| Male | 34/47 (72%) |
| **Manifestation** |  |
| Skin | 25/47 (53%) |
| Bone marrow | 18/47 (38%) |
| 0 EN-sites | 4/38 (11%) |
| 1-2 EN sites | 30/38 (79%) |
| > 2 EN sites | 4/38 (11%) |
| **ECOG PS** |  |
| 0-1 | 16/30 (53%) |
| ≥2 | 14/30 (47%) |
| **B-symptoms** |  |
| No | 12/38 (32%) |
| Yes | 26/38 (68%) |
| **LDH** |  |
| Normal | 9/30 (30%) |
| Elevated | 21/30 (70%) |
| **Immunohistochemistry** |  |
| BPDCN-specific | 47/47 (100%) |
| CD56<sup>+</sup> | 47/47 (100%) |
| CD123<sup>+</sup> | 46/47 (98%) |
| TCL1 | 40/47 (85%) |
| Immature lineage marker | 37/47 (79%) |
| CD34 | 5/47 (11%) |
| TdT<sup>+</sup> | 34/47 (72%) |
| T-lineage markers | 46/47 (98%) |
| CD2<sup>+</sup> | 12/41 (29%) |
| CD3<sup>+</sup> | 8/47 (17%) |
| CD4<sup>+</sup> | 44/47 (94%) |
| CD7<sup>+</sup> | 6/47 (13%) |
| B-lineage marker CD79A<sup>+</sup> | 39/47 (83%) |
| Myeloid-lineage markers | 44/47 (94%) |
| CD33<sup>+</sup> | 43/47 (91%) |
| CD117<sup>+</sup> | 10/47 (21%) |
| MPO<sup>+</sup> | 4/47 (9%) |
| Ki-67 (median, range) | 50% (20 – 90%) |
| **1st line therapy (available in 27 cases)** |  |
| Upfront HSCT (Auto/Allo) | 6 (3/3) (22%) |
| CHOP-like regimen | 10 (37%) |
| ALL regimens* | 5 (19%) |
| Others | 8 (30%) |
| BSC | 4 (15%) |
| Abbreviations: ALL, acute lymphoblastic leukemia; BPDCN, blastic plasmacytoid dendritic cell neoplasm; BSC, best supportive care; CHOP, cyclophosphamide/hydroxydaunorubicin/vincristine/prednisolone; ECOG, Eastern Cooperative Oncology Group; EN, extranodal; HSCT, hematopoietic stem cell transplantation; LDH, lactate dehydrogenase; MPO, myeloperoxidase; yrs, years. |  |
| *GMALL, B-ALL protocol |  |
| **DeVIC protocol (n=2), azazitidine-mono (n=4), radiation therapy (n=1), DHAC (n=1) |  |

### The mutational landscape of BPDCN identified by WES

To comprehensively characterize the mutational landscape in BPDCN patients, WES of patient-derived tumor biopsies was performed with matched normal DNA in three cases. Variant calling for SNVs and indels in individual samples involved filtering strategies to avoid FFPE-derived artefacts and spurious mutations (see **Supplementary methods**). From these mutations the MUTSIGCV algorithm identified 41 significant candidate driver genes (p < 0.001; 20 genes with q < 0.1; 21 additional genes with q < 0.2; **Supplementary Table 4**) ^35^. Following said bioinformatic annotation, putative oncogenic diver mutations were identified in all BPDCN cases. In total 13,908 presumably deleterious mutations, affecting 4,507 genes were observed. SNVs and indels comprised 39.8% of these mutations (5,532), of which 4,524 were missense (81.8%), 289 nonsense (5.2%) and 468 indel mutations (8.5%). Mutations affecting splice sites represented 4.4% of estimated somatic mutations and 0.2% of the observed mutations were non-stop mutations. At an intermediate-low median tumor mutational burden (TMB) of 2.57 mutations/Mbase (range 0.52 – 10.21), we observed micro-satellite-instability (MSI)-related hypermutation in five cases, which significantly exceeded expectations for an acute hematological malignancy ^36^. TMB did not differ between tumor-only samples and samples with matched normal tissue (Wilcoxon test p = 0.0708).

Across the entire cohort, *TET2* (Tet Methylcytosine Dioxygenase 2) was the gene most frequently mutated, followed by *KMT2D* and *EP300*. An oncoplot integrating clinical features and mutational status of all putative oncogenic driver genes according to our MUTSIGCV analysis is provided in **Figure 1A**. The list of significant candidate driver genes included several genes previously implicated in BPDCN and further expanded on these ^5,11,14,18,20,37,38^. Subsequent gene set enrichment analysis delineated a significant impact of oncogenic mutations on the epigenetic regulation of gene expression (95.7% of cases; *TET2, DNMT3A, KMT2D, SETD2, IDH2*), RTK-RAS (93.6%; *NRAS, MET, EGFR*), NOTCH (76.6%; *NOTCH2, CREBBP, EP300*) and WNT signaling pathways (59.6%; *CTNNB1, MED12*) (**Supplementary Figure 1**). Several known or potentially actionable therapeutic vulnerabilities were observed, including recurrently mutated residues in activating receptors (e.g. *EGFR*) and activating GTPases (e.g. *NRAS*) (**Figure 1B, C**) and others ^39,40^. Exemplarily, activating (bioinformatically predicted or functionally predescribed) mutations in tyrosine kinase genes such as *MET, PDGFRA* and methyltransferase enzymes, including *EZH2* or deleterious mutations in TSGs like *CDKN2A*, pose viable targets for molecularly tailored therapy approaches (e.g. capmatinib, avapritinib, tazemetostat, palbociclib), particularly in the clinical setting of a molecular tumor board, as high levels of evidence suggest therapeutic efficacy across a variety of cancers^41-44^. Functional implications of all reported mutations according to the CADD score are summarized in **Supplementary Table 5** alongside annotation for affected protein domains due to SNVs and indels. Among MUTSIGCV-selected genes a median CADD score of 25.1 (mean 25.9) was observed, further underscoring pathobiological relevance of our observations^45^.

**Figure 1.**
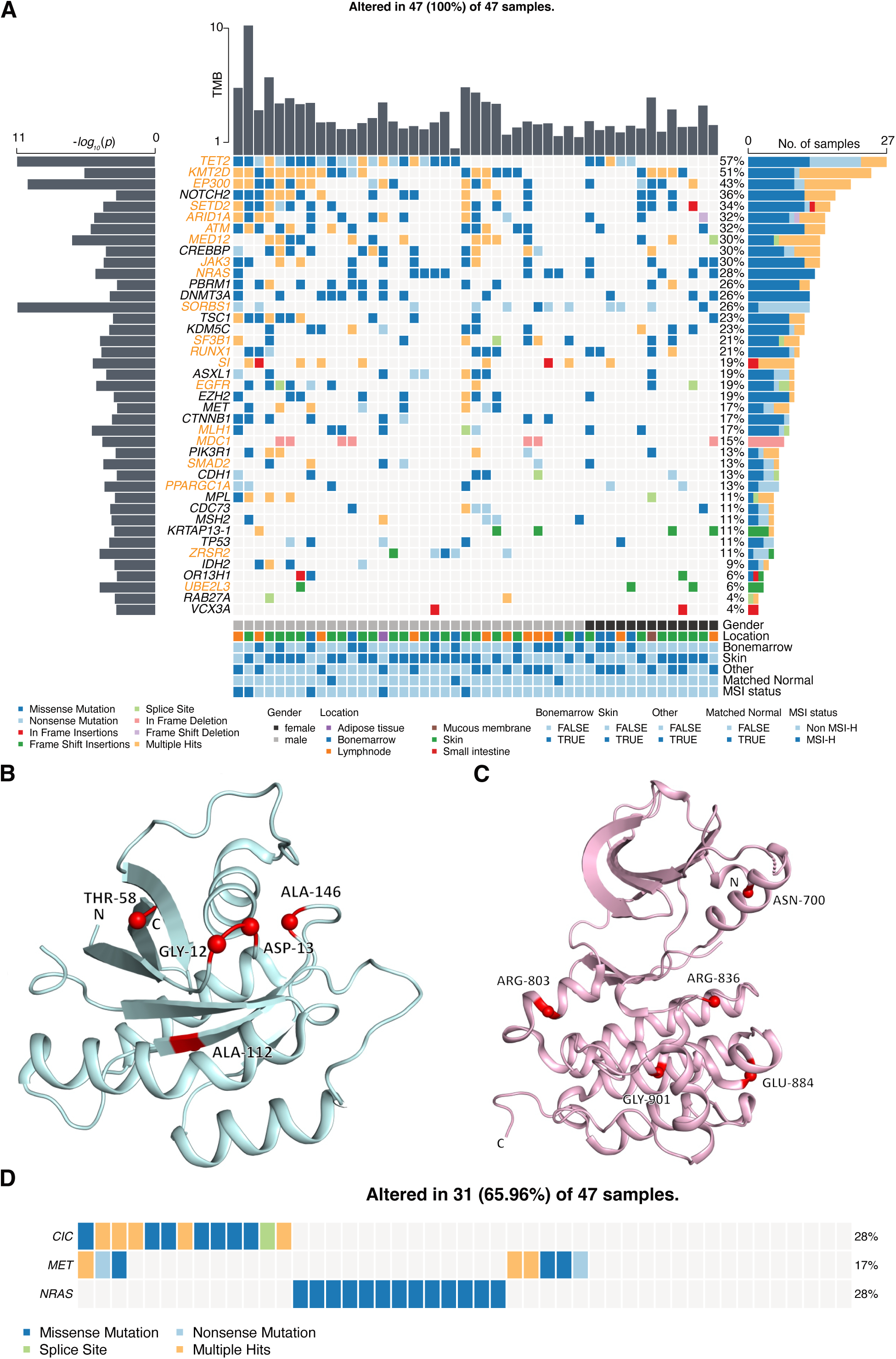
Mutational landscape in BPDCN. (A) Oncoplot showing potential driver genes inferred by MUTSIGCV with tumor mutational burden (TMB; upper bar plot), *-log*_*10*_-transformed p-vales (left bar; orange gene names q < 0.1, black gene names p < 0.001) and number of affected samples (right bar). Mutation types are color coded, and covariates are shown below for each sample (covariate ‘Other’ refers to samples with tissue affected other than skin or bone marrow). (B) Spatial clustering of mutations within the protein structure of NRAS (PDB ID 6E6H) and (C) eGFR (PDB ID 6TG0). (D) Mutual exclusive occurrence of *NRAS* with *CIC* and *MET*; type of mutation is color coded.

A pair-wise Fisher’s exact test for mutual exclusivity or co-occurrence of mutations, found evidence for mutual exclusivity in *CIC* and *MET* correlated with *NRAS* mutations (**Figure 1D**). Moreover, several, statistically significant combinations of co-occurrences were observed, including *MPL* and *NOTCH2, SETD2*, and *TSC1* as well as *EGFR* and *EP300* alongside *SETD2* (**Supplementary Figure 2**).

### Impact of molecular subtype and gender on significantly mutated driver genes and mutational relatedness with neighboring entities

Next, we asked if there was an association between certain significantly mutated driver genes and molecular or clinical subtypes of BPDCN. First, we observed a significant enrichment of mutations affecting *ARID1A, ATRX*, and *CTNNB1* in MSI^high^ cases, which is in keeping with previous reports in colorectal, gastric and endometroid cancer and others^46-48^. *ARID1A* poses a prominent interaction partner of mismatch repair (MMR) protein MSH2, while *ATRX* mutations were previously found to be a sex-biased predictor of MSI-status across a variety of solid tumors and *CTNNB1* was recurrently implicated in the pathogenesis of an especially unfavorable subset of endometroid tumors and in immune checkpoint inhibitor resistance^47,49,50^ (**Figure 2A**).

**Figure 2.**
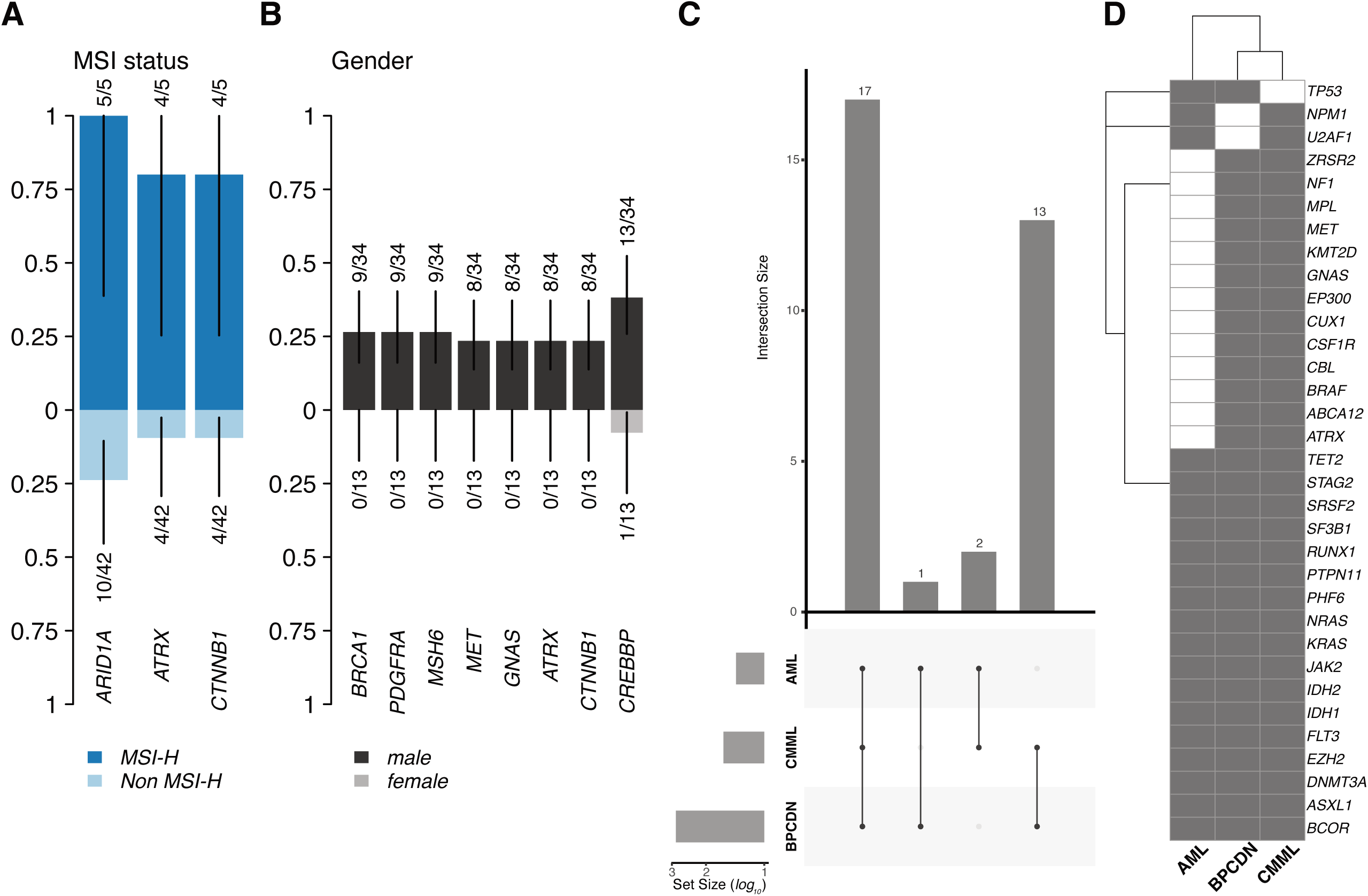
Significantly enriched genes and comparison to CMML/AML. Genes significantly enriched in MSI-H samples (A) and with a male mutation bias (B); number of affected samples and total number of samples are given and scale on the y-axis denotes proportion of mutated samples. (C) UpSet plot showing the overlap between BPDCN samples (n = 47, this study, MUTSIGCV genes selected), CMML (n = 76, Papaemmanuil *et al*. 2013, Papaemmanuil *et al*. 2016, Tyner *et al*.) and AML (n = 672, Tyner *et al*.) for genes mutated in at least two samples per data set (only overlapping genes are shown); data for CMML and AML was retrieved via CBIOPORTAL.^*60,62,63*^. (D) Overlapping genes between the three data sets for genes mutated in at least two entities.

Beyond the recently described sex-biased implications of *ZRSR2* mutations in male BPDCN patients, we detected an additional, statistically significant enrichment of mutations affecting *ATRX* and *CTNNB1* alongside other putative oncogenic drivers of BPDCN pathogenesis including *BRCA1, PDGFRA, MSH6, MET, GNAS*, and *CREBBP* in male patients (**Figure 2B**).

To assess the mutational similarity of BPDCN with neighboring entities, we compared our cohort results with samples from AML (n = 672) and CMML (n = 76) patients from the TCGA. Apparently, CMML is most closely related to BPDCN, with shared mutational drivers including *BRAF, CSFR1, EP300, MET*, and *ZRSR2*. In addition, we found several mutations to co-occur between all three entities including *TET2, SRSF2, SF3B1, NRAS, KRAS*, and *IDH1/2*. Merely *TP53* mutations were found to be an exclusive significant commonality between AML and BPDCN, potentially reflecting the shared, aggressive nature of the two diseases (**Figure 2C,D**).

### Recurrent copy number alterations in BPDCN

Recurrent somatic copy number alterations (SCNAs) in BPDCN were identified from the OncoScan CNV. We found 5 recurrent regions classified as pathogenic (e.g. del3p21.31 resulting in a loss of *SETD2* in 11 patients and del10q23.2 resulting in a loss of *PTEN* in five patients) and 12 regions as likely pathogenic according to X-CNV. Pathogenic or likely pathogenic regions were significantly enriched in regions of loss (**χ**^2^ test p = 4.0×10^−4^). We were hereby able to retrieve previous observations from conventional cytogenetic studies regarding recurrent aberrations affecting 3p21.31, 10q23.2, 12p13.2, 13q34 and 17p (losses) and 3q26.1, 8q24.3 (gains) and others, revealing a characteristic spectrum of cytogenetic aberrations for BPDCN in our cohort. Next, we analyzed the genes affected by these SCNAs and observed recurrent losses of tumor-suppressor genes (TSGs; most of which were found to be biallelic in a subset of patients) such as *CDKN2A, NOTCH1, RB1*, and *BRCA2* and copy-number gains in oncogenes including *IDH2, U2AF1, MET*, and *EZH2* (**Figure 3A-C**). Details on SCNA chromosomal location, affected genes and statistical observations regarding the significance of observed gains and losses are provided in **Supplementary Tables 6 and 7**.

**Figure 3.**
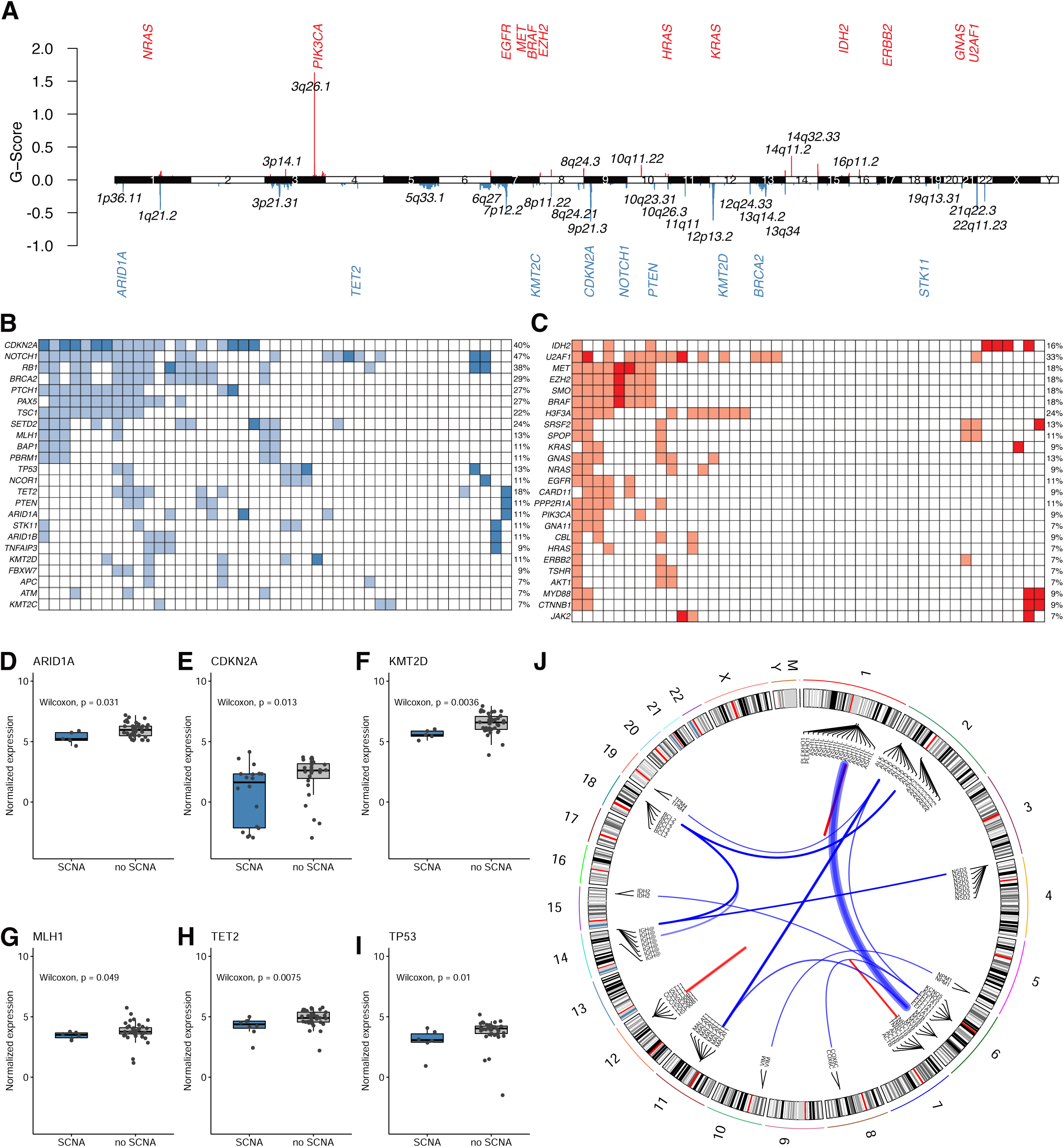
Genomic alterations and gene fusions. (A) Location of SCNAs along the genome and GISTIC G-scores (G = Frequency × Amplitude; red bars denote gains and blue bars losses; gene names refer to affected oncogenes and tumor suppressor genes within identified regions). (B) Tumor suppressor genes affected by copy number losses (light blue: low level deletions; dark blue: high level deletions) and (C) oncogenes affected by copy number gains (light red: low level amplifications; dark red: high level amplifications). Panels (D-I) show significant differences in expression levels of selected genes in regions of copy number losses. (J) Known cancer and *MYB* fusion identified in BPDCN samples with respect to their genomic location; red links indicate intra-chromosomal fusions, blue links indicate inter-chromosomal fusions, respectively. Link width correlates with the number of reads supporting the fusion event.

### Deregulation of PDGFR signaling and ECM interactions in BPDCN by RNA-Seq

Next, gene expression data from bulk BPDCN RNA-Seq were compared with peripheral blood pDCs (CD45^+^ CD123^+^ BDCA2^+^ CD3^−^) from healthy donors, which were available through a previous study ^51^. On average 75.1 million reads (median 82.3) were successfully mapped to the human reference per tumor sample and 29.6 million reads (median 30.4) per normal sample, respectively. Differential gene expression analysis unveiled results similar to those obtained by Togami *et al*., including upregulation of *BCL2, MYB*, and others ^11^. Given the more comprehensive cohort analyzed in the current project and reflecting the molecular heterogeneity of the disease, we here found an upregulation of oncogenes such as *PDGFRA/B, EGFR, FGFR1*, and others as well as a down-regulation of inflammatory mediators such as *IL2, IL22*, and *CXCL8*. Despite the results from our MUTSIGCV analysis in which *PDGFRA* mutations were shown to be events of only borderline significance (p = 0.05), we found a pathobiological relevance of these mutations reflected in its simultaneous significant upregulation, contributing to the phenotype of BPDCN. In line with this finding a pathway enrichment on differentially regulated genes between healthy pDC samples and BPDCN identified a tumoral downregulation of metabolic processes implicated in oxidative phosphorylation. PDGF signaling, NCAM1 interactions and cell cycle accelerators were over-expressed. In addition, several processes associated with interactions with the extracellular matrix as a recurrent feature in BPDCN and in concert with previous observations regarding mutational impairment through *IKZF1* inactivation and others were significantly upregulated(**Figure 4 A, B**) ^38,52^.

**Figure 4.**
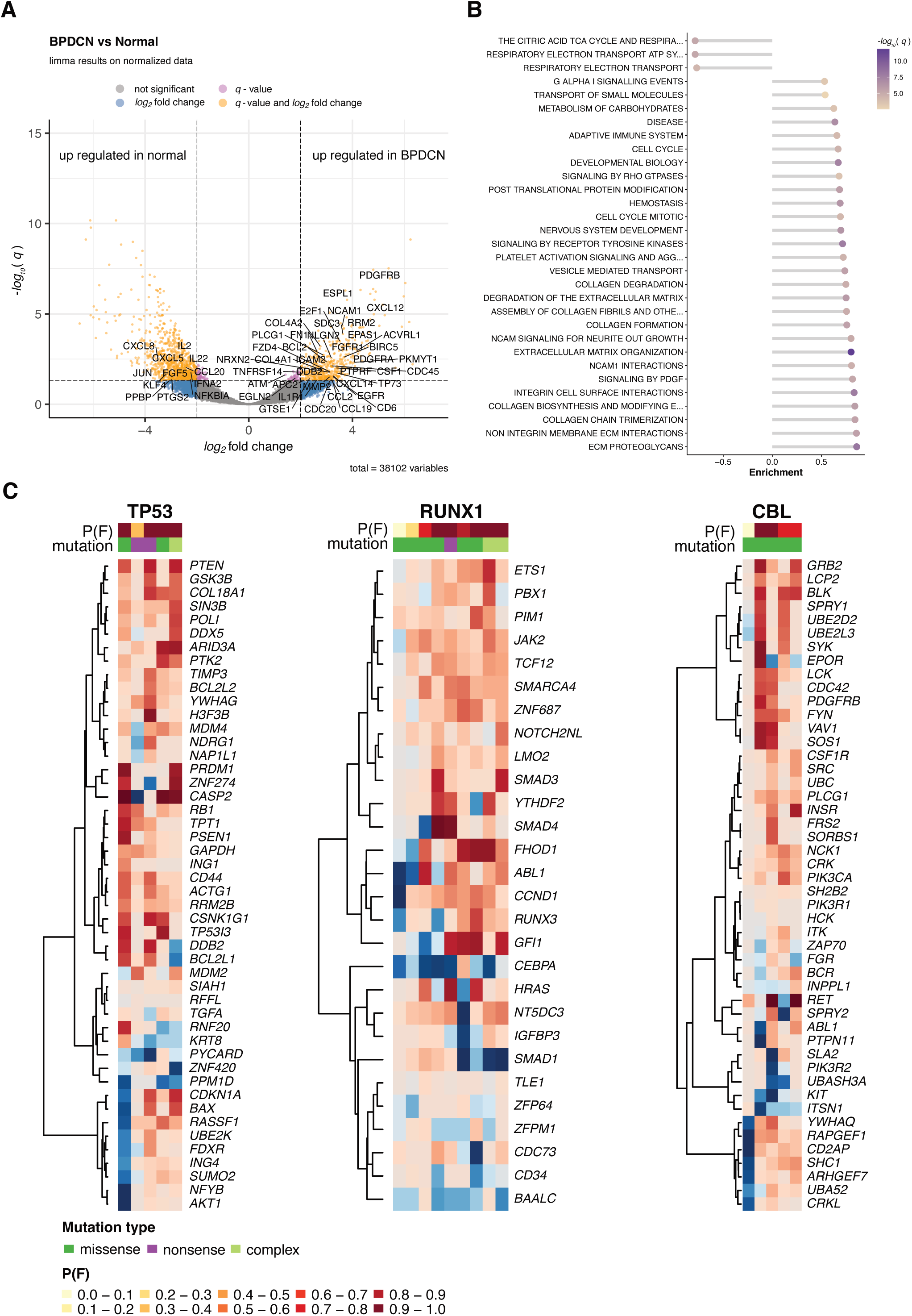
Patterns of differentially expressed genes in BPDCN and *trans* effects of mutations on gene expression. (A) Volcano plot showing *log*_*2*_ fold-changes and *-log*_*10*_ scaled q-values of the expression analysis of BPDCN samples against normal controls (peripheral blood pDCs). Dotted lines denote significance level (q < 0.05) and absolute *log*_*2*_ fold-changes above 2; known tumor suppressor genes, oncogenes and genes related to cytokine-cytokine receptor interaction, cell adhesion molecules cams, cell cycle, P53 signaling pathway and pathways in cancer (all retrieved from KEGG) are highlighted if they are significant. (B) Pathway enrichment analysis against REACTOME gene sets on differentially expressed genes (q < 0.01, absolute *log*_*2*_ fold-change > 2); enrichment scores (length of bar) and *-log*_*10*_ scaled q-values (color-coded dots) are displayed for pathways with q < 0.05 and absolute enrichment scores > 0.5. (C) Significant *trans* effects of mutations on gene expression for *TP53, RUNX1*, and *CBL* identified applying XSEQ; red represents high expression and blue represents low expression; P(F) denote probability values are color coded; type of mutation is color coded.

### Integrated analysis between DNA alterations and RNA expression

Combining the SCNA and RNA-Seq data enabled us to validate the copy-number loss of several TSGs including *CDKN2A, KMT2D*, and *TP53* on the level of gene expression, despite the limited samples sizes and regardless of mutational status (**Figure 3D-I**).

XSEQ integrates mutational and gene expression data to assess the impact of significant gene mutations on RNA-Seq-derived profiles ^53^. This identified “*trans*-effects” with high confidence for *TP53, RUNX1* and *CBL* that hint at the key role of these genes in shaping the malignant transcriptional profile and phenotype in BPDCN. Most prevalently, mutations affecting the transcriptional regulator *RUNX1*, which guides the differentiation of hematological precursor cells, poses a plausible driver for the development of the malignant phenotype of the disease, to which *CBL* as a ubiquitin ligase and *TP53* as a central tumor suppressor gene further contribute (**Figure 4C**).

### Fusion genes

Employing FUSION CATCHER we identified recurrent fusion events in BPDCN. Hereby we observed the previously described *MYB* fusions (**Figure 3J**), which were successfully validated by FISH in all cases (**Supplementary Figure 3**). Further recurrent oncogenic driver fusions, including *BCL2* and *ALK* aberrations, are listed in **Supplementary Table 8**.

### Integrated WES/RNA-Seq/SCNA reveals two mutationally distinct BPDCN subtypes of diverging ontogeny

A deconvolution of bulk gene expression profiles based on single cell transcriptome data allowed to estimate the abundances of cell types within the mixed cell population of our BPDCN biopsies. We focused on the distribution of dendritic cell and monocyte subtypes for each case using signatures from previously published scRNA-Seq data ^30^. Subsequently, hierarchical clustering identified two distinct subpopulations within our cohort, in which plasmacytoid dendritic cells (pDCs), common DCs (DC1 and DC2) and different monocyte subtypes were prevalent at variable frequencies. A typical pDC-derived subtype composed of a relatively pure pDC population (named C1) and an atypical (common)cDC-enriched subtype (named C2). The latter is driven by the overexpression of DC1/2 markers *CLEC9A* and *CD1C* beyond typical pDC markers such as *CLEC4C, GZMB* which shape the C1 phenotype As could be expected from the report by Renosi *et al*., we additionally observed a subset of BPDCN cases enriched in DC5 signatures (AS-DCs) in more than half the cohort samples, which were, however, equally distributed across both clusters ^5^ (**Figure 5A**). While close, and in some cases even clonal, relations between BPDCN and CMML denote a recurrent feature, we observed syn-or metachronous CMMLs or AMLs in six C2 cases and only in two C1 cases. Testing at a cut-off of 65 years or older the C2 cluster was significantly enriched for elderly patients (C1: 10 young and 14 old patients; C2: 3 young and 20 old patients; Fisher exact test p = 0.04899) with a distinct mutational distribution (**Supplementary Figure 4**). Intriguingly, genomic analysis of these newly defined subtypes revealed that C1 patients displayed an enrichment in *EP300, ARID2, NF1, NOTCH2*, and *SF3B1* mutations, whereas atypical C2 cases were enriched for *DNMT3A* and *SRSF2* mutations (**Figure 5B, Supplementary Figure 5A**). Additionally, C1 showed a significantly higher TMB, which is in typical BPDCN (Wilcoxon test p = 0.002; **Figure 5C**). In order to validate these observations, we performed an additional multi-omics factor analysis into which mutational SCNV and gene expression data are integrated unsupervised. Hereby, we observed a strong association in two out of 10 factors that comprised the two novel BPDCN subtypes, being mainly driven by gene expression and somatic mutations (**Figure 5D-I**) ^54^. Gene set enrichment of genes with positive weights on one of the factors revealed an enrichment of formation of the cornified envelope, keratinization, and neutrophil degranulation, whereas negative weighted genes were associated with axon guidance, Fc epsilon receptor signaling, and signaling by EGFR/NGF (REACTOME pathway; q < 0.05; **Supplementary Figure 5B**,**C**). A visual summary of molecular features of BPDCN, derived from this study, alongside their respective functional implications is provided in **Figure 6**.

**Figure 5.**
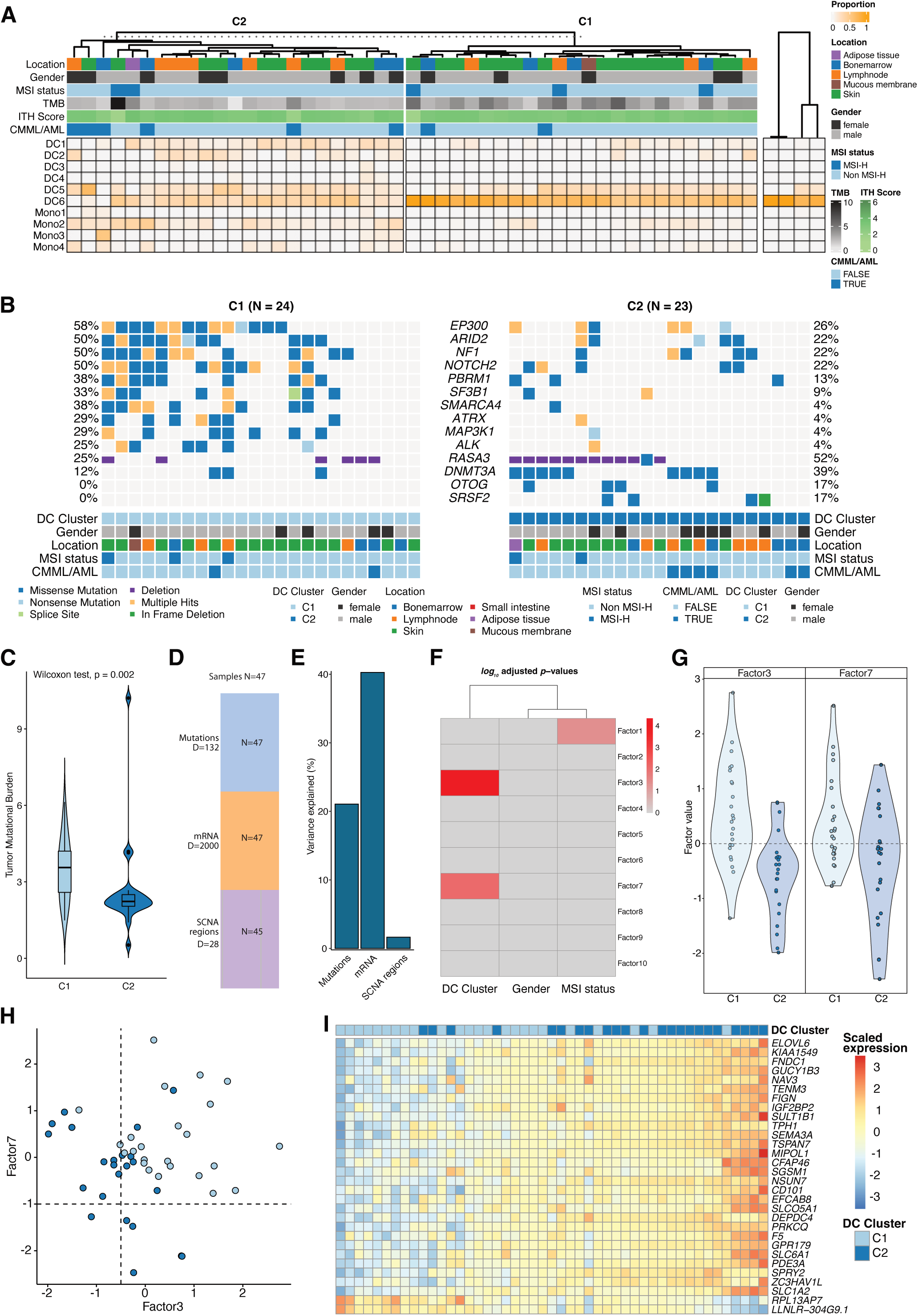
Identification of pDC and cDC-derived subtypes. (A) Proportion of dendritic cells (DC1-DC6) and monocytes (Mono1-Mono4) according to the deconvolution analysis for BPDCN samples (left heatmap with annotations; TMB refers to tumor mutational burden and ITH score describes the inferred intra-tumor heterogeneity) and normal controls (peripheral blood pDCs; shown in the right heatmap without additional annotations). The optimal number of clusters was inferred using hierarchical clustering and average silhouette method. (B) Co-oncoplot of genes identified as significantly enriched between the two cluster (see **Supplementary Figure 5A** for details). (C) Tumor mutational burden estimates for each cluster. Panel (D) shows the number of samples (‘N’) and features per feature group (‘D’) used in the multi-Omics factor analysis (MOFA+). (E) Variance explained per feature after training MOFA+ (Mutations = SNVs and indels; mRNA = normalized expression values; SCNA region = genomic location of somatic copy number alterations). (F) Correlation of identified factors with selected covariates (only correlations where p_adj_ < 0.05 are shown). (G) Beeswarm plots of latent Factor3 and Factor7 for each dendritic cell cluster. (H) Scatter plot of estimated factor values for each sample; light blue refers to C1 and dark blue to C2. (I) Scaled gene expression values of top genes (n=20) that correlated with Factor3; cluster annotation for each sample is shown above each sample.

**Figure 6.**
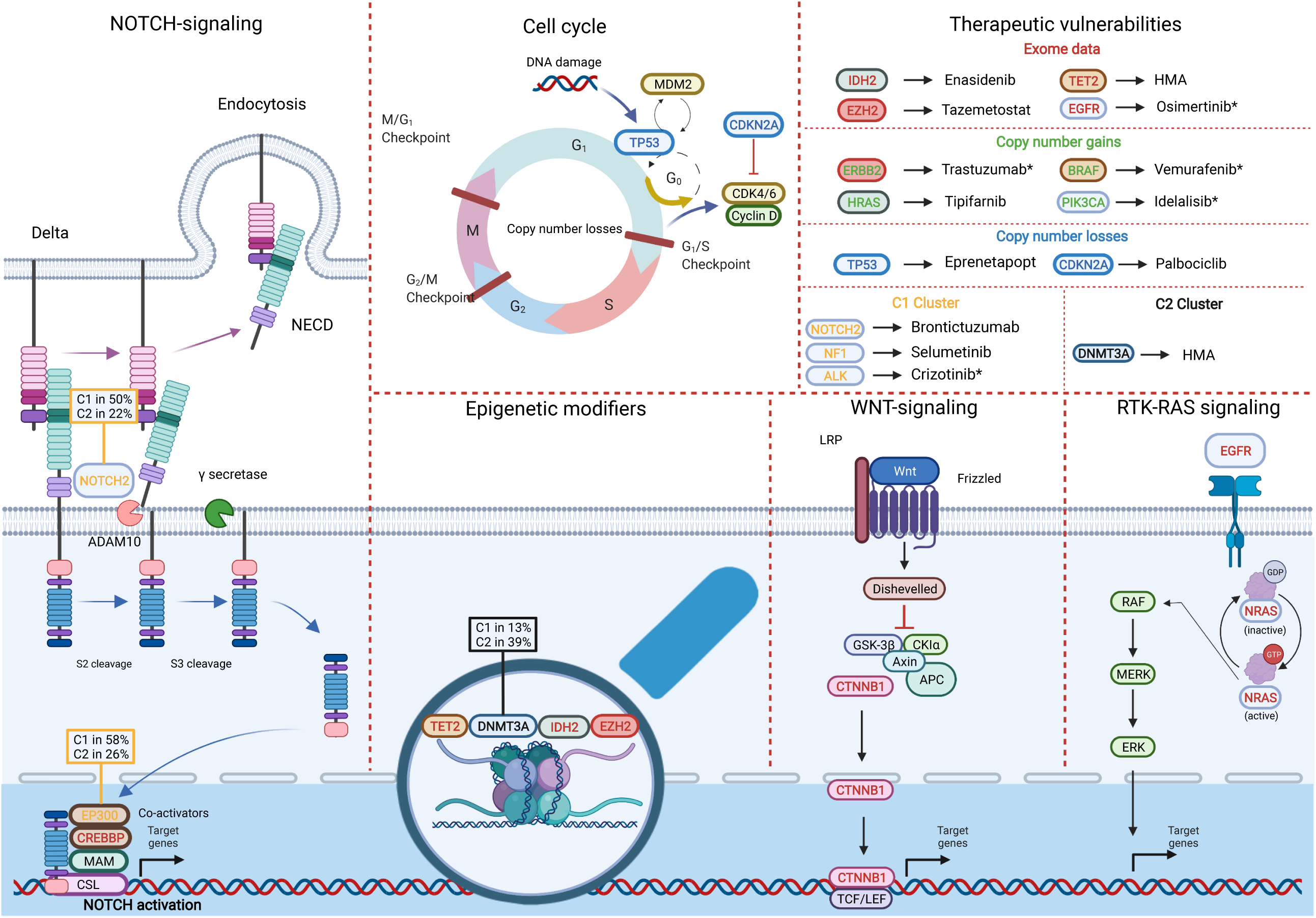
Visualization of predominantly altered pathways and potential therapeutic targets in BPDCN. Whole exome sequencing (WES) revealed frequent alterations NOTCH (*NOTCH2, CREBBP, EP300*), WNT (*CTNNB1*) and RTK-RAS (*EGFR, NRAS*) signaling as well as in epigenetic modifiers (*TET2, DNMT3A, EZH2* and *IDH2*). Additionally, SCNA analysis identified two copy number losses (*TP53* and *CDKN2A*) significantly affecting cell cycle regulation towards cancer progression. Integrated WES/RNA-Seq analysis divided two distinct BPDCN subtypes in accordance with a subtype-specific mutational signature. The C1 cluster harbours *NOTCH2, EP300, NF1* and *ALK* mutations whereas *DNMT3A* mutations were found to be recurrent features within the C2 cluster. Further, therapeutically targetable vulnerabilities uncovered by comprehensive genomic profiling are summarized.

## Discussion

In this hitherto most comprehensive, paired genomic and transcriptomic study of BPDCN, supplemented by CNA analysis, we systematically defined recurrent mutations, CNAs, and gene fusions alongside predominant mutation and transcriptional signatures beyond the scope of previous efforts ^5,11-14,18,51,52,55,56^. The present study hereby made three novel and essential observations.

Firstly, a precision oncology roadmap of targetable alterations and vulnerabilities is provided. We describe a substantial subgroup of patients with MSI^high^, offering a strong rational for an immune checkpoint inhibitor treatment ^57^. This is further supported by the recurrent deletion, in concert with significantly reduced expression of *MLH1*, as a prominent MMR gene and deleterious mutations of *MSH6* ^58,59^. Confirming and expanding on results from previous studies, an enrichment in mutations altering the epigenetic regulation of gene expression was observed (e.g. *TET2, EZH2*) alongside recurrent mutations in various members of the RTK-RAS and NOTCH signaling pathways, with *TET2* and *KMT2D* being the most frequently altered genes in our cohort. The concept of pathobiological significance and a crucial role of these alterations in BPDCN is further supported by recurrent deletions. Oncogenic mutations and significantly induced expression of tyrosine kinases like *PDGFRA* and others such as CDK-affecting mutations/deletions pose alternative prominent potential targets for therapeutic intervention ^44^. Unlocking BPDCN for precision oncology efforts is an essential step forward in the treatment of this disease, as a substantial subset of patients does not qualify for curative treatment approaches due to age and frailty ^3^.

Secondly, we extend the understanding of the BPDCN mutational landscape which is significantly shaped by gender and molecular subgroup (MSI^high^ vs. MSS) alongside the molecular taxonomy within the continuum of CMML, AML and BPDCN. Regarding the mutational frequencies in BPDCN and its neighboring entities, we provide further evidence of the close molecular relatedness between BPDCN and CMML, while the biology of BPDCN appears to be distinct from AML, not merely on the transcriptional level or by immunophenotype but also with regard to the genomic landscape, as we observed only *TP53* mutations as an exclusive shared feature between AML and BPDCN ^60-62^. The latter, in concert with mutations in *RUNX1* and *CBL* were further shown to impact gene expression profiles extensively through significant and biologically meaningful “trans-effects” and thereby shaping the malignant phenotype in a substantial subset of patients. This phenomenon not only highlights the need for integrative genomics approaches in the characterization of the pathobiological basis of rare cancers, but also underscores the regulatory impact of significantly mutated genes on altered signaling and ultimately the malignant phenotype of BPDCN in particular.

Thirdly, through the deconvolution of bulk RNA-Seq data, we identified two transcriptionally defined subsets of BPDCN, independent from localization or tumor cell content/purity within the sample. By estimating the abundances of dendritic cells and monocyte subtypes for each case the cellular origin of the predominant malignant BPDCN clone was investigated. Hereby, we discovered two distinct subpopulations within our cohort, a typical pDC-derived subtype composed of a practically pure pDC population (C1) and an atypical (common)cDC-enriched subtype (C2), in which both pDCs, cDCs and different monocyte subtypes were prevalent at variable frequencies. Next, we identified distinctive features of these newly defined subtypes within the genomic data. An integrated analysis of RNA-Seq and WES revealed a significantly higher TMB in typical BPDCN (C1) cases alongside a distinct mutational profile as C1 patients displayed an enrichment in *EP300, ARID2, NF1, NOTCH2*, and *SF3B1* mutations, whereas atypical C2 cases were enriched for *DNMT3A* and *SRSF2* mutations. Intriguingly, these distinct features were reflected within the clinical presentation of these subgroups, with C1 patients being significantly younger and C2 patients showing a trend towards inferior survival, albeit in a limited subset of patients with sufficient available follow-up data (**Supplementary Figure 6**). In basic concordance with our results, Renosi *et al*. recently demonstrated the transcriptional heterogeneity of BPDCN regarding the occurrence of both a canonical pDC phenotype in one and an AXL^+^ SIGLEC6^+^ DC (AS-DC)-related profile in the other subset ^5^. In our analysis, we find these cases to be equally distributed among both typical BPDCN cases as well as the atypical cDC-derived variant.

Future studies are now aimed at more comprehensive cohorts of patients with complete clinical follow-up and available samples of both tumor and, if available, unaffected blood or bone marrow samples in order to investigate the relationship of our multi-omics defined BPDCN subtypes with the emerging role of clonal hematopoiesis in BPDCN ^20^.

In conclusion, our findings of several promising targets and molecular signatures for precision oncology approaches alongside a distinct mutational landscape, divergent from AML and strong relatedness with CMML coupled with the identification and genomic characterization of two transcriptionally and clinically distinct subsets within the spectrum of BPDCN significantly advance the understanding of this entity. Further, these observations underscore the potential clinical utility of mutational and transcriptional testing in order to refine the classification and ultimately treatment allocation in DC- and monocyte-derived malignancies.

## Supporting information

Pathway-enrichment analysis

Combinations of mutational (exclusive) co-occurrences

FISH validation of two representative cases harboring a MYB fusion according to RNA-Seq

Age dependent mutated genes

(A) DC-Cluster dependent distribution of mutations. (B) Feature set enrichment of positive and (C) of negative weights for Factor3

(A) Overall survival by Kaplan-Meier and (B) Hazard-ratio according to DC-Cluster

MUTSIGCV analysis. Separate Excel file

Identified variants per sample and predicted functional impact

SCNA results by GISTIC analysis (losses) and functional impact assessment

SCNA results by GISTIC analysis (gains) and functional impact assessment

Fusions

## Data Availability

Bam files from WXS and raw fastq files from RNA-Seq have been deposited in the European genome-phenome archive (EGA) under the accession number EGAS00001006166. OncoScan Array data has been deposited in Gene Expression Omnibus (GEO) under accession number GSE200113.

https://ega-archive.org

https://www.ncbi.nlm.nih.gov/geo/

## Declarations

### Ethics approval and consent to participate

This retrospective study was approved by the ethics committee of the University of Lübeck (reference-no 18-311) and conducted in accordance with the declaration of Helsinki. Patients at the Reference center for Hematopathology have provided written informed consent regarding routine diagnostic and academic assessment, including genomic studies of their biopsy specimen alongside transfer of their clinical data.

## Author contributions

Study concept: NG, ACF, HM

Data collection: NG, AK, JS, HW, SS, KN, KK, VB, HM, SS, EM

Data analysis and creation of figures and tables: AK, NG, HW, VB, HB, NvB

Initial Draft of manuscript: NG.

Critical revision and approval of final version: all authors.

## Acknowledgements

The authors would like to thank Tanja Oeltermann for her skilled technical assistance. AK and HB acknowledge computational support from the OMICS compute cluster at the University of Lübeck.

