## Supplementary Material for "Integrative molecular profiling identifies two molecularly and clinically distinct subtypes of blastic plasmacytoid dendritic cell neoplasm"

**Supplementary Materials and methods.**

**Clinicopathological assessment**

Patients at the Reference center for Hematopathology provided written informed consent regarding routine diagnostic and academic assessment, including genomic studies of their biopsy. Reassessment of histopathological diagnosis, according to the current WHO classification criteria identified 74 cases of BPDCN from which 47 cases with sufficient FFPE tissue samples for subsequent molecular analysis were selected and subjected to a comprehensive immunophenotypic workup. Antibodies and positivity cut-offs employed in the current study are summarized in **Supplementary Table 1**.

**Exome data and Variant Calling**

Coverage per sample was estimated using mosdepth (v0.3.2)^1^ and matched tumor-normal pairs were verified applying BAM-matcher^2^.

Variant calling was performed following GATKs best practices for somatic variant calling (matched normal-tumor mode for samples with normal tissue available and tumor-only mode for samples without normal tissue available). Briefly, Mutect2 (GATK)^3^ was applied to all *BAM* file with gnomAD variants as germline resource and the b38 exome panel from the 1000 genome project as a panel of normal, capturing the expected repertoire of germline variants to be expected in a Central European study population. Next, FFPE read orientation artifacts were identified and removed according to GATK guidelines. Filtered variants were annotated using Variant Effect Predictor (VEP v103, GRCh38; adding CADD v1.6, dbNSFP v4.1a^4^, and gnomAD r3.0 as additional annotations)^5^ and annotations were converted into *MAF* format using vcf2maf (v1.6.21) (doi:10.5281/zenodo.593251); coverage was extracted directly from the vcf INFO field. The top 20 frequently mutated genes (FLAGS)^6^ were removed from further analysis and remaining somatic variants were filtered as follows: minimum coverage of 40, minimum alternative allele coverage of 5, minimum variant allele frequency of 10%, and only variants with a frequency < 0.1% in 1000 genomes, gnomAD, or ExAC were considered for subsequent downstream analysis. High impact variants (CADD score > 10) in tumor suppressor and oncogenes according to Vogelstein *et al.* were filtered as such that a minimum coverage of 20 minimum, minimum alternative coverage and, and minimum variant allele frequency of 10% was required^7^. Genes mutated more often than expected were identified applying MutSigCV (v1.41)^8^ and potential drivers were identified suing p < 0.001.

**RNA-Seq data**

Gene expression profiles were normalized applying MIXnorm (v0.0.0.9000; 50 iteration, tolerance set to 0.1)^9^, which removes unwanted biological and technical effects from formalin-fixed paraffin-embedded (FFPE) material that can bias the signal of interest.

Pathway enrichment analysis against REACTOME gene sets (MSIGDF R package v7.2) on significantly different expressed genes was performed using a rank-MANOVA based approach as implemented in MITCH (v1.6.0; priority on significance)38; NF-𝜅β pathway (KEGG) was added manually to the REACTOME gene set.

Identified fusions were filtered for known cancer and *MYB* fusions to remove potential false positives following the FusionCatcher guidelines. Additionally, fusion only detected by BOWTIE were discarded. Fusions were visualized using chimeraviz (v1.20.0)^10^.

**Statistical analysis**

For analysis and visualization the following R packages were applied: Tidyverse (v1.3.1)^11^ for data handling; maftools (v2.10.05)^12^ to summarize, analyze, and visualize variant data; EnhancedVolcano (v1.12.0) (https://github.com/kevinblighe/EnhancedVolcano) to show the relationship between significance and fold-changes; ComplexHeatmap (v2.10.0) and pheatmap (v1.0.12) to draw heatmaps; ggpubr (v0.4.0) for box and violin-plots; UpSetR (v1.4.0) for UpSet plots.

Progression-free survival and overall survival (PFS, OS) were calculated from the date of diagnosis and censored at last clinical contact. Survival (PFS and OS) according to potential prognostic factors was estimated by means of the Kaplan–Meier method and univariate log-rank test. Additionally, hazard ratios were calculated using a Cox proportional hazards regression model. Survival analysis was carried out employing the R packages survival (3.3-1) and survminer (v0.4.9).

Microsatellite instability was estimated with MSIseq (v1.0.0)^13^ using a custom made database of mono repeats GRCh38 (*find.mono.repeats* function) and intra-tumor heterogeneity was estimated on the entropy of somatic mutation (mDITHER-score) using DITHER (v1.0)^14^. The functional impact of mutations was assessed using xseq (v0.2.2)^15^; to estimate *trans* effects, potential *cis* effects of somatic copy number alterations were removed.

An unsupervised framework (Multi-Omics Factor Analysis v2, MOFA2 v1.4.0)^16^ was applied to integrate the different omics data sets (WES data, RNA-Seq data, SCNA data) to be able to discover principal sources of variation in the data. Briefly, mutated genes were considered if they were mutated in at least 4 samples selected, normalized expression values of the top 2,000 most variable genes were selected and all SCNA regions were considered for training. Feature set enrichment analysis was performed against REACTOME gene sets using parametric *t*-test on gene expression data and Benjamini-Hochberg correction to adjust p-values for multiple testing.

**Supplementary Table 1.** Antibodies and conditions used throughout the study

| **Antibody** | **Manufacturer** | **Clone** | **Dilution** | **Incubation period** |
| --- | --- | --- | --- | --- |
| CD3 | Leica | LN10 | RTU | 30min |
| CD4 | Leica | 4B12 | RTU | 30min |
| CD33 | Leica | PWS44 | RTU | 30min |
| CD34 | Leica | Qdend | RTU | 30min |
| CD56 | Leica | CD564 | RTU | 30min |
| CD79a | Leica | JCB117 | RTU | 30min |
| CD117 | leica | EP10 | RTU | 30min |
| CD123 | Leica | BR4MS | 1:100 | 30min |
| Lysozym | DAKO Agilent | A 0099 | 1:10000 | 30min |
| Ki67 | Leica | K2 | RTU | 30min |
| MPO | Leica | 59A5 | RTU | 30min |
| CD068 | DAKO Agilent | PG-M1 | 1:100 | 30min |
| TCL-1 | Menarini | MRQ-7 | 1:100 | 30min |
| Tdt | Leica | SEN28 | RTU | 30min |
| RTU, ready to use dilution | | | | |

**Supplementary Table 2.** Confirmatory Sanger and amplicon-based next generation sequencing of selected variants

| **CaseID** | **Chromosome** | **Reference_Allele** | **Alt_Allele** | **Hugo_Symbol** | **CDS_position** | **AAChange** | **vaf** | **AAPos** | **Methode** |
| --- | --- | --- | --- | --- | --- | --- | --- | --- | --- |
| BPDCN_05 | chr17 | C | T | TP53 | 524 | TP53:p.R175H | 0,4746 | 175 | NGS |
| BPDCN_10 | chr17 | T | C | TP53 | 488 | TP53:p.Y163C | 0,3872 | 163 | NGS |
| BPDCN_15 | chr21 | C | A | RUNX1 | 611 | RUNX1:p.R204L | 0,5082 | 204 | NGS |
| BPDCN_21 | chr4 | G | A | TET2 | 3886 | TET2:p.G1296R | 0,4364 | 1296 | NGS |
| BPDCN_25 | chr2 | G | A | IDH1 | 394 | IDH1:p.R132C | 0,4636 | 132 | NGS |
| BPDCN_26 | chr4 | C | T | TET2 | 1858 | TET2:p.Q620* | 0,3874 | 620 | Sanger |
| BPDCN_45 | chr4 | A | T | TET2 | 2314 | TET2:p.K772* | 0,2741 | 772 | Sanger |
| BPDCN_49 | chr20 | C | T | ASXL1 | 1762 | ASXL1:p.Q588* | 0,3182 | 588 | Sanger |
| BPDCN_49 | chr4 | C | T | TET2 | 2431 | TET2:p.Q811* | 0,2698 | 811 | Sanger |

**Supplementary Table 3.** Immunohistochemical profile of the study cohort

| **Case IDN-Nr.** | **localization** | **CD3** | **CD4** | **CD33** | **CD34** | **CD56** | **CD79a** | **CD117** | **CD123** | **ASD** | **Lyso** | **Ki67** | **MPO** | **PGM1** | **TCL1** | **TdT** |
| --- | --- | --- | --- | --- | --- | --- | --- | --- | --- | --- | --- | --- | --- | --- | --- | --- |
| BPDCN_01 | lymph node | - | + | +/- | - | + | - | - | + | - | - | 60% | - | - | + | +/- |
| BPDCN_02 | skin | +/- | + | + | - | + | +/- | - | + | - | - | 30% | - | +/- | + | +/- |
| BPDCN_05 | bone marrow | - | + | - | - | + | - | - | + | - | - | 90% | - | - | + | - |
| BPDCN_06 | lymph node | - | - | + | - | + | +/- | +/- | + | - | - | 60% | - | - | + | + |
| BPDCN_07 | skin | - | + | +/- | - | + | - | - | + | - | - | 60% | - | - | + | + |
| BPDCN_08 | skin | - | +/- | +/- | - | + | +/- | - | + | - | - | 30% | - | +/- | + | + |
| BPDCN_09 | skin | - | + | +/- | - | + | + | - | + | - | - | 80% | - | - | +/- | +/- |
| BPDCN_10 | lymph node | - | - | + | - | + | +/- | +/- | + | - | - | 25% | - | - | + | +/- |
| BPDCN_11 | skin | - | + | + | +/- | +/- | +/- | - | + | - | - | 30% | - | + | + | + |
| BPDCN_12 | bone marrow | - | + | +/- | - | + | +/- | - | + | - | - | 30% | - | +/- | + | +/- |
| BPDCN_13 | skin | - | + | + | - | + | - | - | + | - | - | 20% | - | - | + | - |
| BPDCN_14 | skin | - | + | + | - | + | - | - | + | - | - | 50% | - | - | + | +/- |
| BPDCN_15 | lymph node | - | +/- | +/- | +/- | +/- | - | +/- | + | - | - | 40% | +/- | - | - | +/- |
| BPDCN_16 | skin | - | + | + | - | + | - | - | + | - | - | 50% | - | + | - | - |
| BPDCN_17 | skin | - | + | +/- | - | + | +/- | - | + | - | - | 50% | - | - | + | + |
| BPDCN_18 | skin | - | +/- | + | - | + | +/- | - | + | - | - | 70% | - | +/- | + | - |
| BPDCN_19 | fatty tissue | +/- | + | - | - | + | + | - | - | - | - | 90% | - | - | - | - |
| BPDCN_20 | skin | - | + | + | - | + | +/- | - | + | - | - | 30% | - | - | + | +/- |
| BPDCN_21 | skin | - | + | +/- | - | + | +/- | - | + | - | - | 30% | - | - | + | +/- |
| BPDCN_22 | lymph node | - | +/- | +/- | - | + | +/- | - | + | - | - | 30% | - | - | + | - |
| BPDCN_23 | skin | - | + | +/- | - | + | + | - | + | - | - | 30% | - | +/- | + | + |
| BPDCN_24 | skin | +/- | + | - | - | + | +/- | - | + | - | - | 70% | - | - | + | +/- |
| BPDCN_25 | lymph node | +/- | - | + | + | +/- | + | +/- | + | - | +/- | 60% | +/- | +/- | - | +/- |
| BPDCN_26 | skin | - | + | + | - | + | + | - | + | - | - | 30% | - | - | + | +/- |
| BPDCN_27 | skin | - | +/- | + | - | + | + | - | + | +/- | + | 60% | - | + | - | - |
| BPDCN_28 | mucosa | - | +/- | +/- | - | + | +/- | - | + | - | - | 60% | - | - | + | + |
| BPDCN_29 | lymph node | - | + | +/- | - | + | +/- | +/- | + | - | - | 30% | +/- | - | + | + |
| BPDCN_30 | lymph node | - | + | + | - | + | + | +/- | + | - | +/- | 60% | - | + | + | +/- |
| BPDCN_31 | skin/mucosa | - | + | + | - | + | +/- | +/- | + | - | - | 60% | - | - | +/- | - |
| BPDCN_32 | lymph node | - | +/- | +/- | - | + | +/- | +/- | + | - | +/- | 30% | - | - | + | +/- |
| BPDCN_33 | skin | - | + | +/- | - | + | +/- | - | + | - | - | 30% | - | - | + | - |
| BPDCN_34 | bone marrow | - | + | +/- | - | + | +/- | +/- | + | - | - | 40% | - | + | + | +/- |
| BPDCN_36 | skin | - | +/- | + | - | + | +/- | +/- | + | - | +/- | 30% | - | - | + | + |
| BPDCN_37 | bone marrow | - | + | - | + | + | - | - | + | +/- | + | 60% | +/- | + | - | - |
| BPDCN_38 | bone marrow | +/- | + | +/- | - | + | + | - | + | - | - | 60% | - | - | + | - |
| BPDCN_40 | bone marrow | - | + | +/- | - | + | + | - | + | - | - | 80% | - | - | +/- | - |
| BPDCN_41 | skin | - | + | + | - | + | +/- | - | + | - | - | 70% | - | +/- | + | - |
| BPDCN_42 | lymph node | - | + | + | - | + | +/- | - | + | - | - | 60% | - | +/- | + | - |
| BPDCN_43 | bone marrow | +/- | + | + | - | + | +/- | - | + | - | - | 50% | - | - | + | +/- |
| BPDCN_44 | bone marrow | - | + | + | - | + | +/- | - | + | - | - | 60% | - | - | + | + |
| BPDCN_45 | bone marrow | +/- | +/- | + | + | + | +/- | +/- | + | - | - | 50% | - | + | - | - |
| BPDCN_46 | skin | - | + | + | - | + | + | - | + | - | - | 30% | - | - | + | + |
| BPDCN_47 | skin | - | + | + | - | + | +/- | - | + | - | - | 40% | - | - | + | +/- |
| BPDCN_48 | lymph node | - | + | + | - | + | + | - | + | - | - | 30% | - | + | + | +/- |
| BPDCN_49 | skin | - | + | + | - | + | + | - | + | - | - | 70% | - | + | + | +/- |
| BPDCN_50 | skin | - | + | + | - | + | +/- | - | + | - | - | 80% | - | - | + | +/- |
| BPDCN_51 | skin | +/- | + | + | - | + | +/- | - | + | - | - | 50% | - | + | + | +/- |
| +; > 60 % of nucleated cells/surface area. +/-; 10 - 60 % of nucleated cells/surface area. -; < 10% of nucleated cells/surface area | | | | | | | | | | | | | | | | |

**Supplementary Table 4.** MutSigCV analysis. Separate Excel file

**Supplementary Table 5.** Identified variants per sample and predicted functional impact. Separate Excel file

**Supplementary Table 6.** SCNA results by gistic analysis (losses) and functional impact assessment. Separate Excel file.

**Supplementary Table 7.** SCNA results by GISTIC analysis (gains) and functional impact assessment. Separate Excel file.

**Supplementary Table 8.** Fusions. Separate Excel file.

**Supplementary Figures**

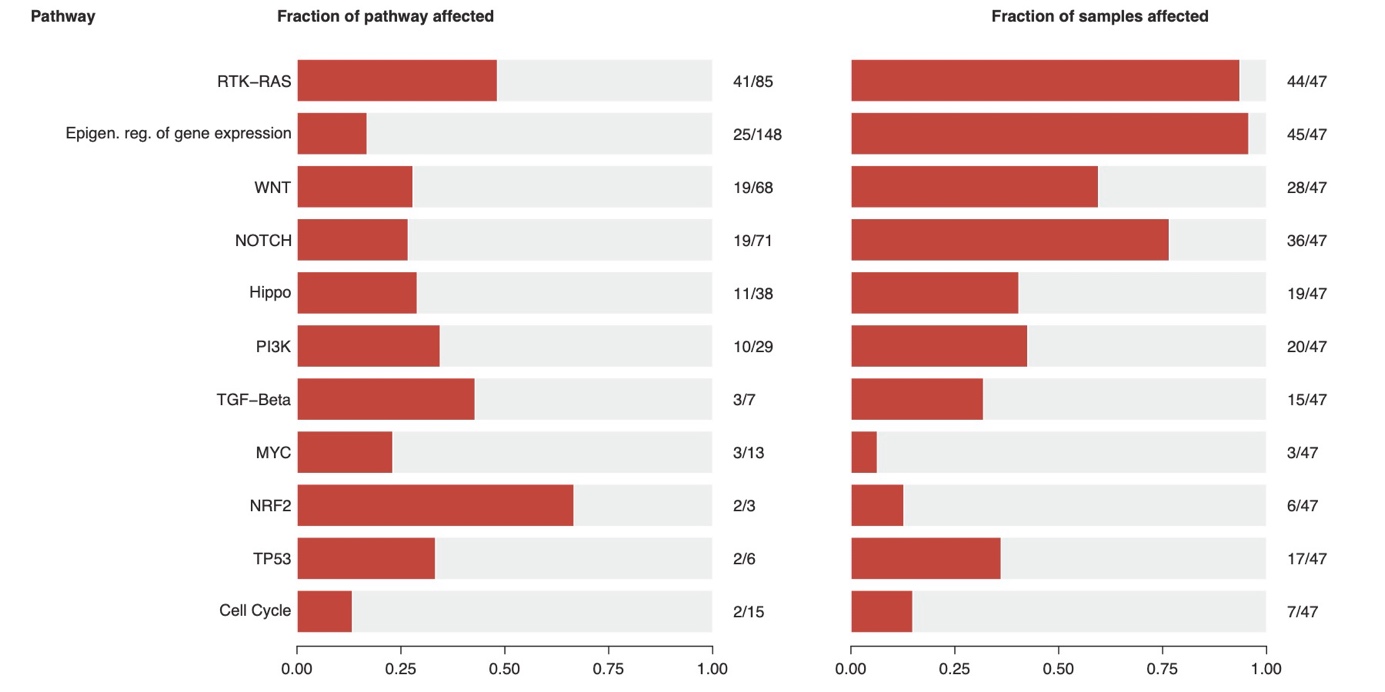

**Supplementary Figure 1.** Pathway-enrichment analysis.

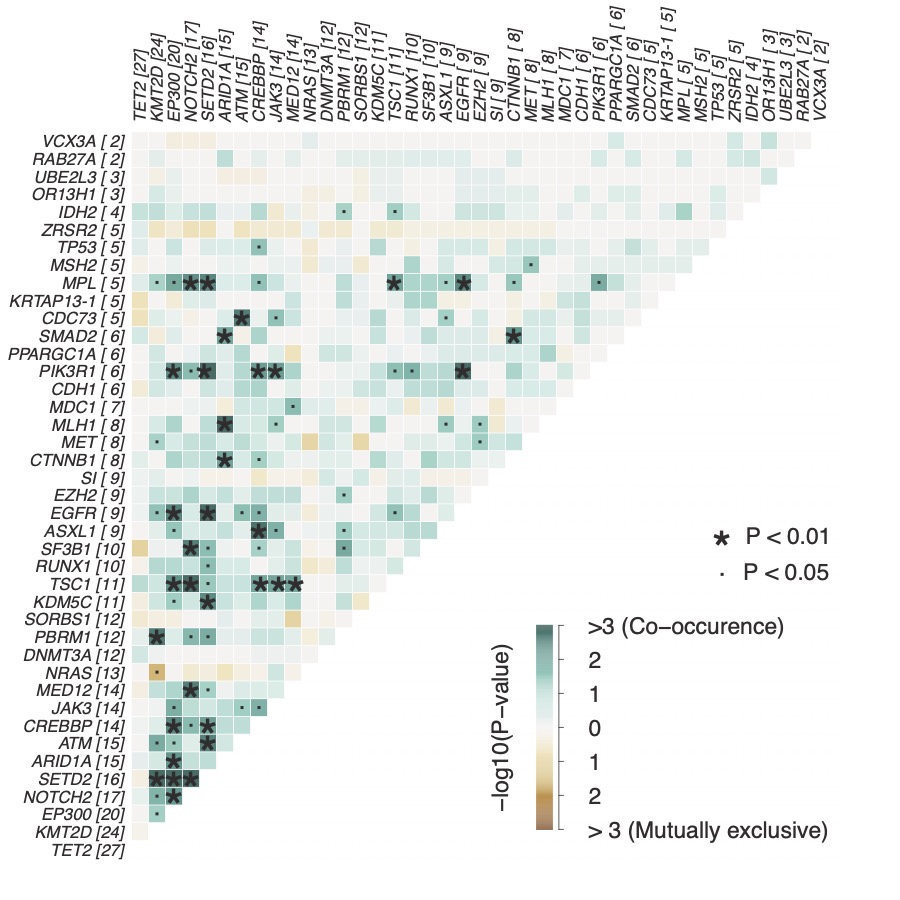

**Supplementary Figure 2.** Combinations of mutational (exclusive) co-occurrences

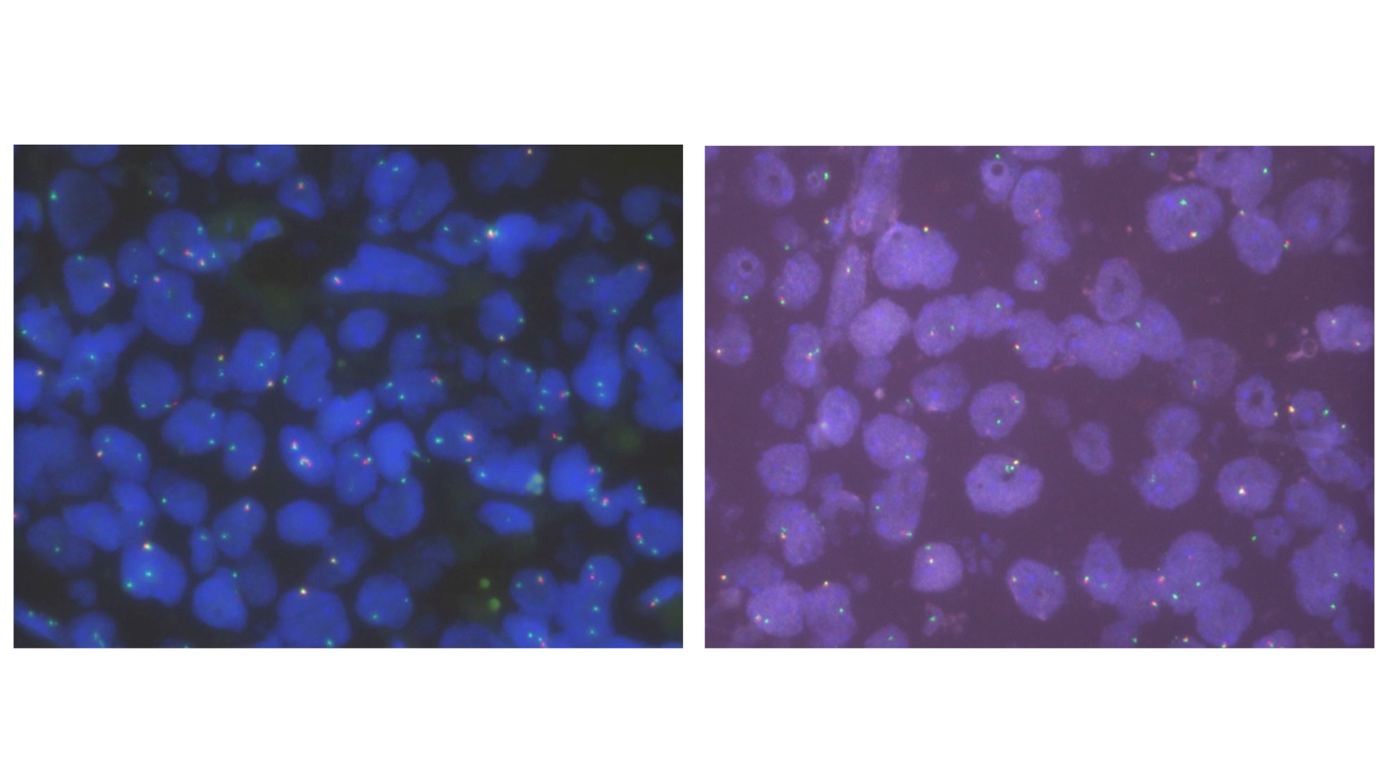

**Supplementary Figure 3.** FISH validation of two representative cases harboring a *MYB* fusion according to RNA-Seq.

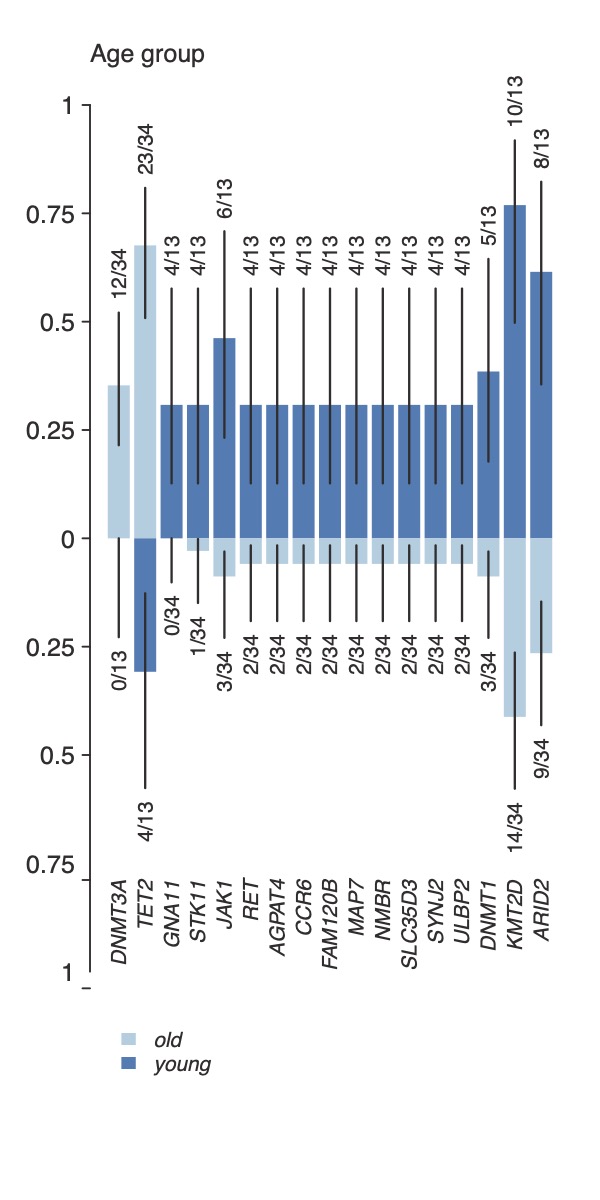

**Supplementary Figure 4.** Age dependent mutated genes.

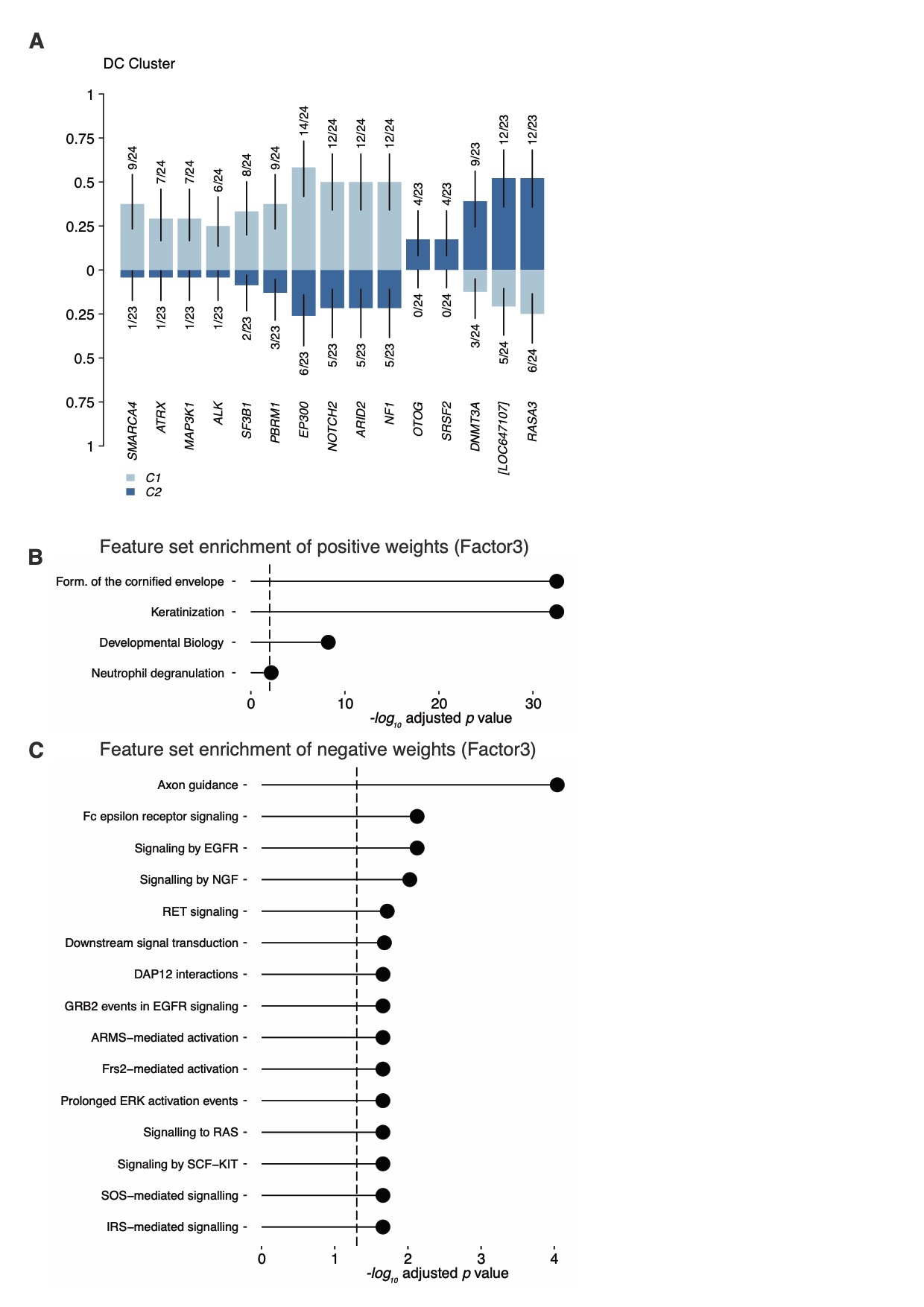

**Supplementary Figure 5.** (A) DC-Cluster dependent distribution of mutations. (B) Feature set enrichment of positive and (C) of negative weights for Factor3.

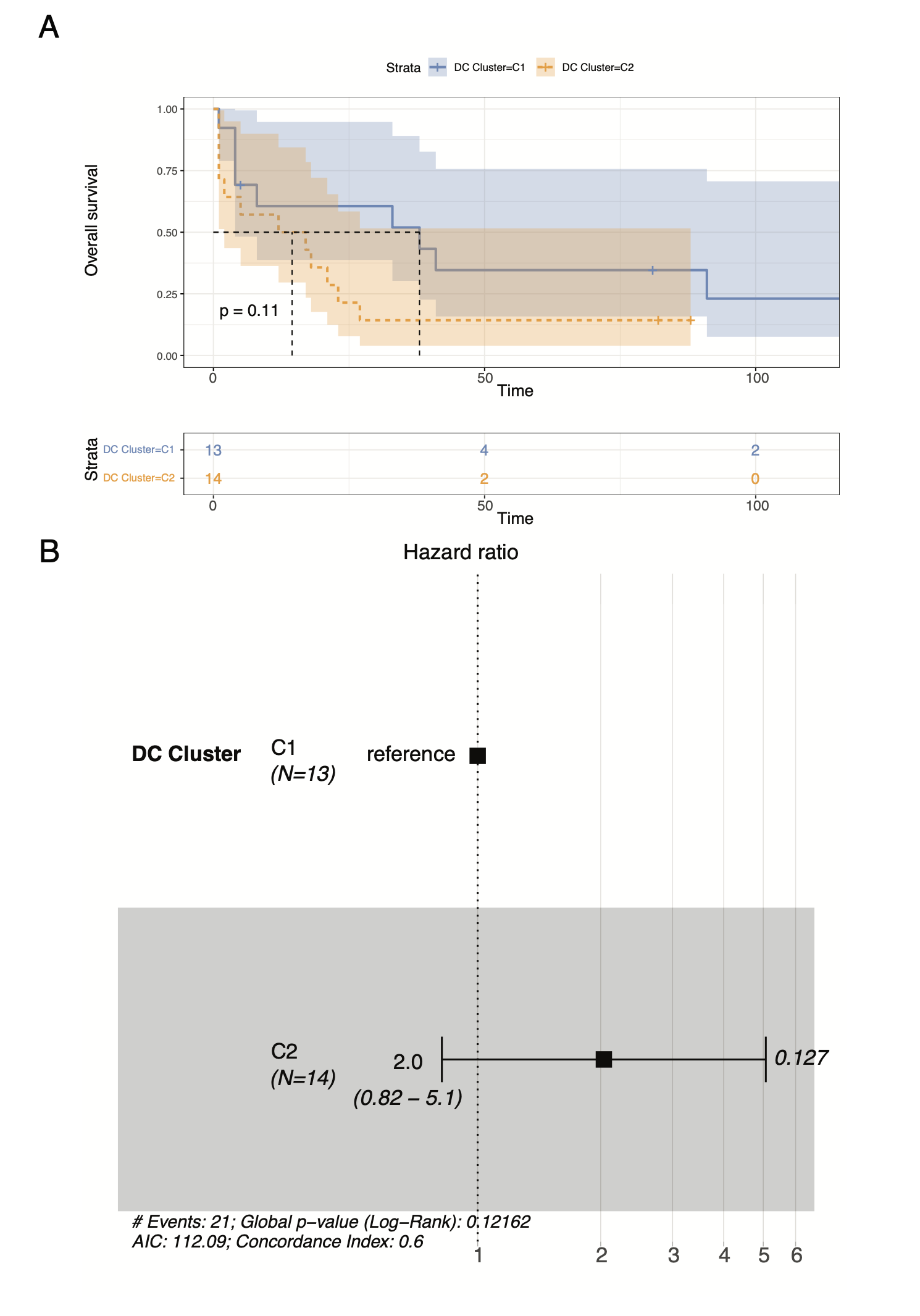

**Supplementary Figure 6.** (A) Overall survival by Kaplan-Meier and (B) Hazard-ratio according to DC-Cluster.
