## Supplementary figures and images for "Integrative molecular profiling identifies two molecularly and clinically distinct subtypes of blastic plasmacytoid dendritic cell neoplasm"

### (A) DC-Cluster dependent distribution of mutations. (B) Feature set enrichment of positive and (C) of negative weights for Factor3

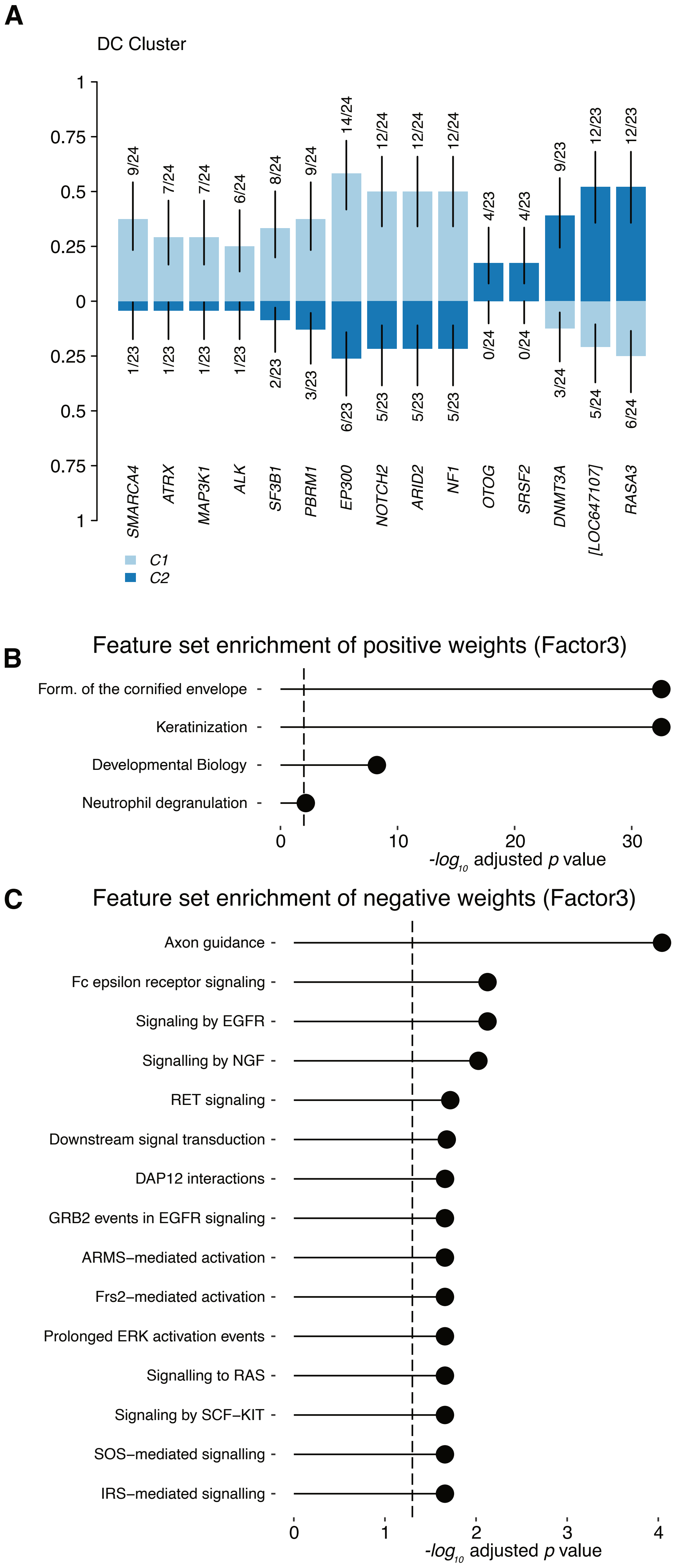

### (A) Overall survival by Kaplan-Meier and (B) Hazard-ratio according to DC-Cluster

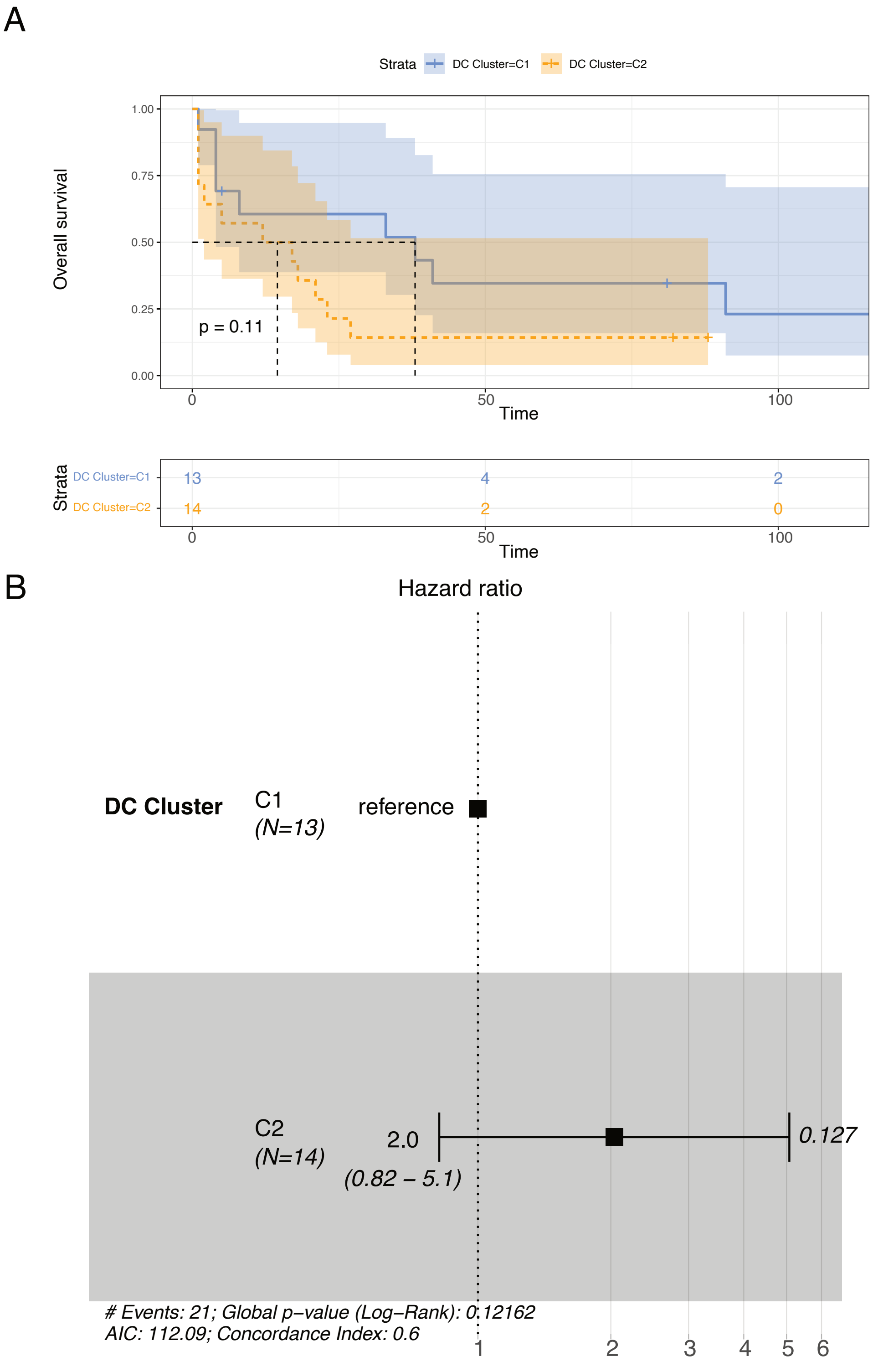

### Age dependent mutated genes

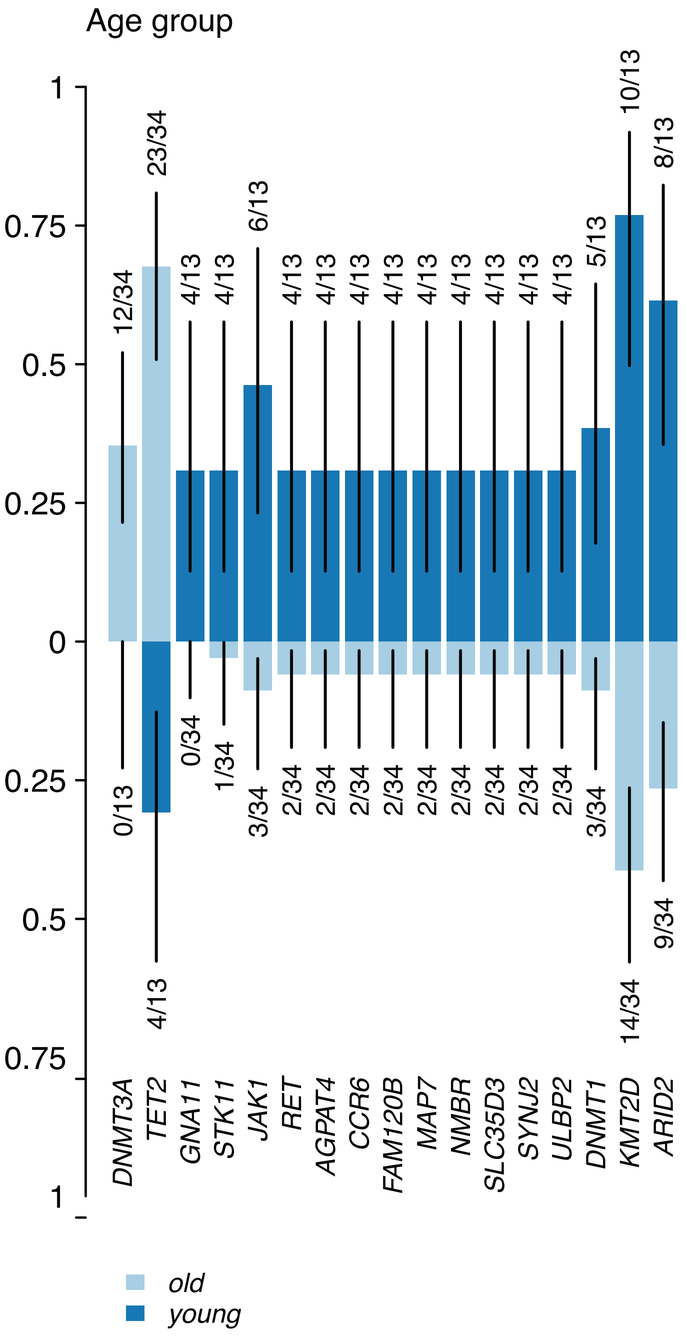

### Combinations of mutational (exclusive) co-occurrences

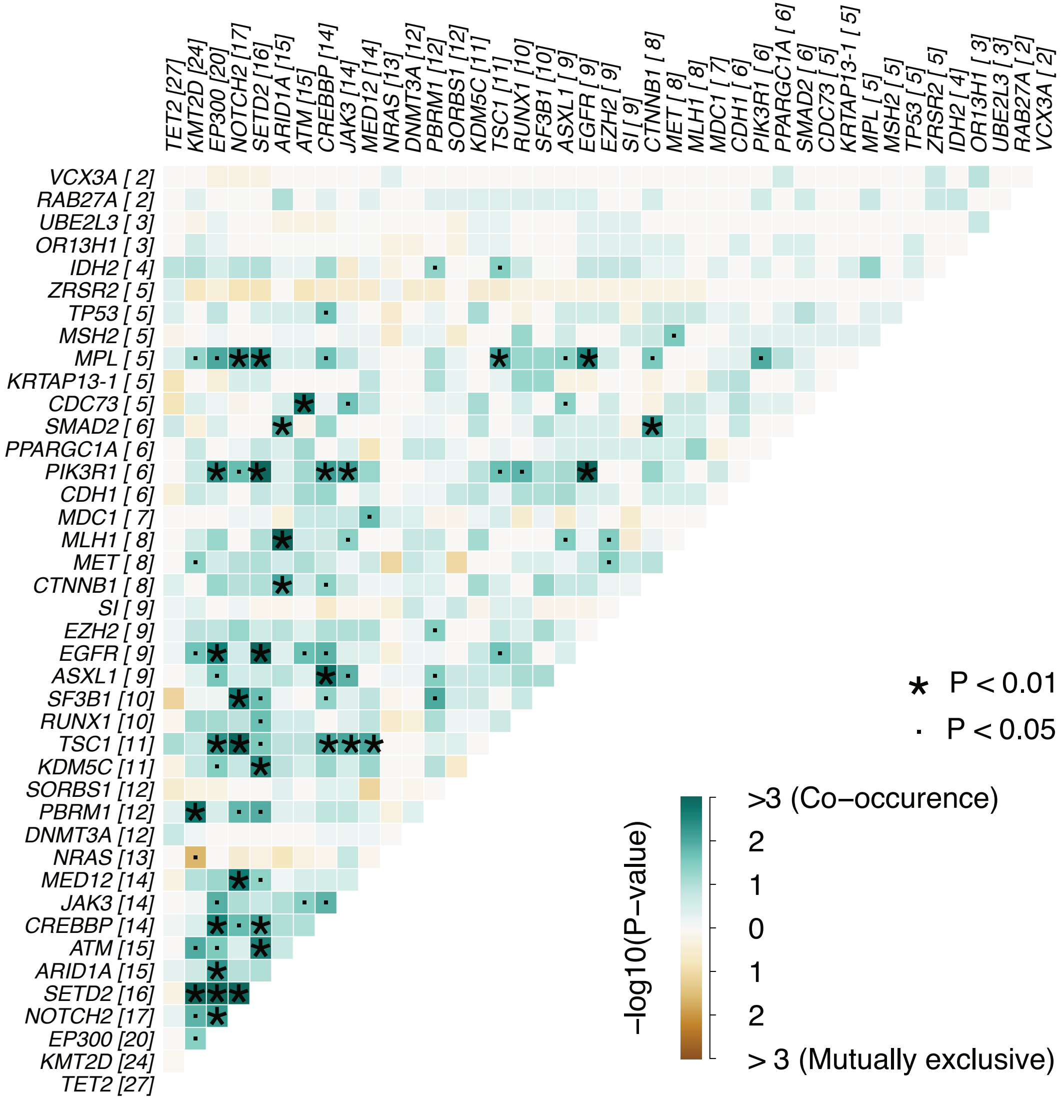

### FISH validation of two representative cases harboring a MYB fusion according to RNA-Seq

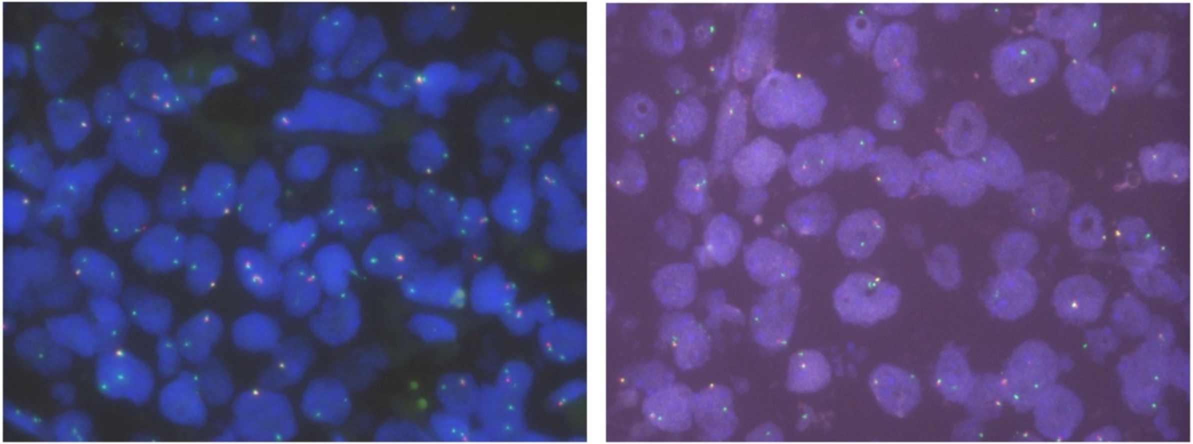

### Pathway-enrichment analysis

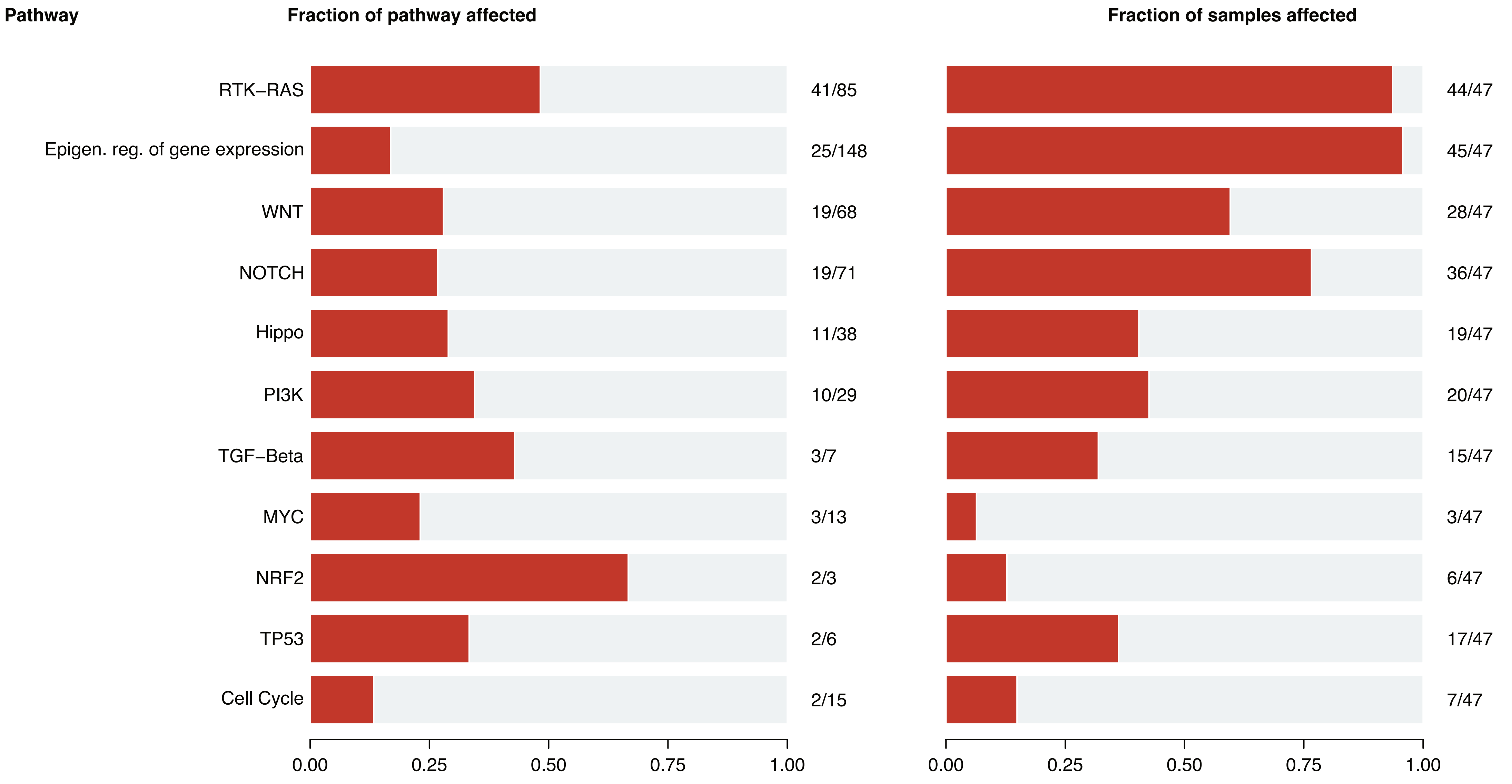
